# Gene mutant dosage is associated with prognosis and metastatic tropism in 60,000 clinical cancer samples

**DOI:** 10.1101/2024.05.13.24307238

**Authors:** Nicola Calonaci, Eriseld Krasniqi, Daniel Colic, Stefano Scalera, Giorgia Gandolfi, Salvatore Milite, Konstantin Bräutigam, Andrea Sottoriva, Trevor A. Graham, Leonardo Egidi, Biagio Ricciuti, Marcello Maugeri-Saccà, Giulio Caravagna

## Abstract

The intricate interplay between somatic mutations and copy number alterations critically influences tumour evolution and patient prognosis. However, traditional genomic analyses often treat these alterations independently, overlooking gene mutant dosage – a key emergent property of their interaction. Here, we develop an innovative computational framework that infers mutation copy number and multiplicity directly from clinical targeted sequencing panels without requiring matched normal samples. Using this approach, we derived gene mutant dosage statistics for over 500,000 mutations across 60,000 clinical samples spanning 39 cancer types. By stratifying more than 20,000 patients according to mutant dosage across multiple oncogenes and tumour suppressor genes, we identified 46 tumour type–specific biomarkers predictive of overall survival. Notably, 13 of these biomarkers across 12 tumour types were undetectable using standard binary mutant/wild-type models. Additionally, 26 biomarkers were recurrently associated with metastatic spread in 10 tumour types, and 20 predicted organ-specific metastatic tropism in 5 tumour types. Alongside confirming known roles for established oncogenes and tumour suppressors, our method reveals, for the first time, gene mutant dosage patterns as independent predictors of prognosis, metastatic potential, and site-specific dissemination across diverse solid tumours. This augmented insight into genomic drivers enhances our understanding of cancer progression and metastasis, holding the potential to foster biomarker discovery significantly.

## 1. Introduction

Cancerogenesis is a complex, multi-step process involving various genetic and epigenetic changes in tumour suppressor genes (TSGs) and oncogenes^1^. Central to this process are somatic mutations and copy number alterations (CNAs), a byproduct of genomic instability, which confer a proliferative advantage to specific cancer cell subpopulations^2^, leading to the dominance of particular clones^3–6^. Genetic biomarkers derived from such driver mutations^7–9^ have significantly impacted our understanding of cancer and its treatment, providing insights into cancer prognosis^10^, and guiding treatment strategies against specific biological vulnerabilities. Despite several successes, however, understanding the prognostic and predictive implications of somatic mutations in key genes remains a challenge^11^, promoting research towards other approaches such as that focus on epigenomic alterations^12,13^. The current approach to biomarker detection via clinical sequencing lacks an integrative view of the tumour genome. Since mutation-derived biomarkers are usually investigated in isolation, the intricate interplay and epistasis within cancer genotypes where these alterations co-occur is potentially overlooked (Figure 1a). A clear example is the recent discovery of gene mutant dosage as a prognostic maker of pancreatic adenocarcinoma^14–16^. This statistic reports the number of copies of a mutation in the genome, and can be computed only by integrating mutation and CNA data. The derivation of potent biomarkers from clinical sequencing is also complicated by other factors. For example, the number of samples available for every combination of tumour type, clinical condition and treatment regimen is still limited, given the high levels of tumour heterogeneity observed in patients^17^. Moreover, the detection of CNAs from clinical targeted sequencing assays is complicated for experimental reasons such as the frequent lack of matched normal samples^18–21^. However, understanding and improving the detection of genetic events that can be exploited in the clinical setting remains crucial, prompting a re-evaluation of current tools and data, especially in light of the recent availability of large clinical cohorts of samples analysed for mutations in hundreds of cancer-related genes through targeted sequencing^22,23^. In this work, we developed INCOMMON, an open-source Bayesian inference tool to determine gene mutant dosage, as well as mutation copy number and multiplicity and the mutant fraction statistics, for a set of mutations detected in a tumour-only clinical targeted sequencing assay. These statistics naturally reveal the joint effect of mutations and focal aneuploidy on TSG inactivation or oncogene activation, and are readily available for integrated biomarker discovery. To process targeted data efficiently, INCOMMON uses Bayesian priors derived from large-scale whole-genome sequencing (WGS) cohorts, together with a likelihood function for read-count data and bulk sample purity. In order to facilitate the usage of orthogonal metrics reported by pathologists in clinical tissue records, INCOMMON models the sample purity probabilistically, resisting potential discrepancies between histological and molecular purity estimates. Compared to state-of-the-art tools like ABSOLUTE^18^, FACETS^19^, PureCN^20^, and CopyWriter^21^, INCOMMON can work without raw data (FASTq/BAM files or pre-segmented depth ratios) or matched normal, facilitating its clinical implementation while lifting limitations of accessing large controlled-access datasets. We used INCOMMON to determine the patterns of co-occurrence of mutations and CNAs from over 500,000 mutations detected in 39 principal solid cancer types of over 60,000 clinical cancer samples from the MSK-MetTropism^24^ (MSK) and AACR GENIE DFCI^25^ (DFCI) cohorts. By unravelling gene mutant dosage statistics from large-scale clinical cohorts, we identified (i) 46 tumour-type specific biomarkers prognostic of overall survival across 12 tumour types (9 hotspot mutations), (ii) 26 frequently associated with metastatic spread in 10 tumour types (5 hotspot mutations) and (iii) 20 that predict organotropism in 5 tumour types (2 hotspot mutations). Besides confirming established results regarding known TSGs and oncogenes, for the first time, our method reveals specific gene mutant dosage patterns as independent predictors of patient prognosis, metastatic propensity, and organ-specific metastatic spread across a wide range of solid tumours.

**Figure 1.**
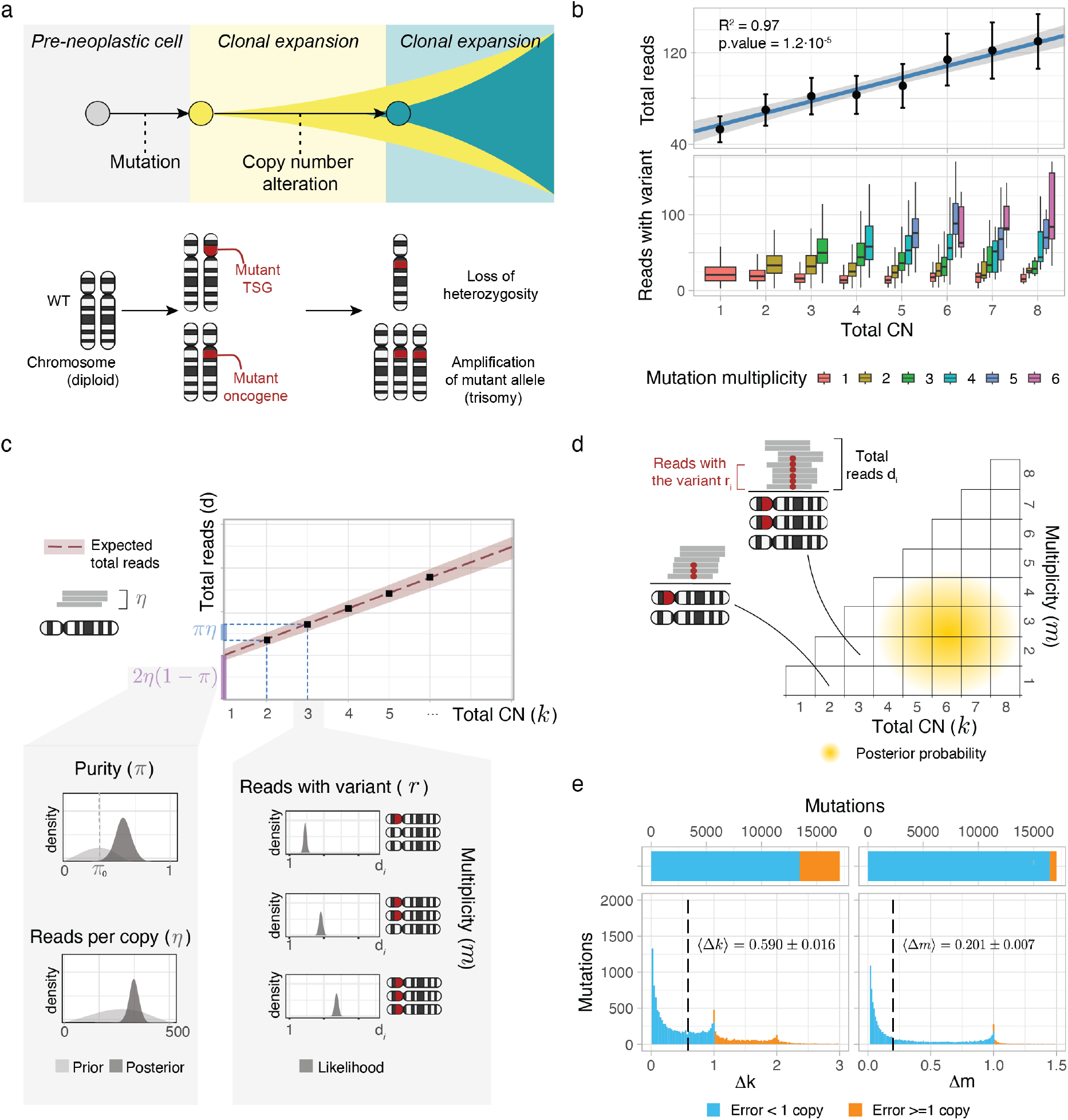
Bayesian inference of mutation multiplicity and copy number with INCOMMON. a. Clonal expansions from a normal cell that acquires first a mutation in a tumour suppressor gene (TSG) or an oncogene, then a copy number alteration (CNA) on the mutant locus, pictured as a loss of the wild-type (WT) allele or a gain of the mutant. b. Linear correlation between total read counts and mutation copy number (CN, top panel), and distribution of reads with the variants (bottom panel) for increasing mutation multiplicity values, for *n* = 8, 339 high-resolution WGS samples available from the PCAWG, HMF and TCGA cohorts. c. INCOMMON is a Bayesian model that leverages a biology-informed prior and infers the posterior distribution of two sample-level parameters (tumour purity *π* and the rate of reads per chromosome copy *η*) and two mutation level parameters (total copy number *k* and mutation multiplicity *m*) from the number of reads with the variant *r* and the total number of reads *d* of each mutation in the sample. d. INCOMMON assigns a posterior probability to any possible configuration of a mutant gene, determined by the co-occurrence of mutations and copy number alterations. e. Performance of INCOMMON across *n* = 50 cross-validation iterations of random-split cross-validation (70% training, 30% test) using ground truth data from PCAWG^**Aaltonen2020**^ and HMF^26^. The deviation from true values of *k* and *m*, Δ*k* and Δ*m*, was less than one copy on average, occurring in 78.6% ± 0.9% of *k* predictions and 96.4% ± 0.4% of *m* predictions.

## 2. Results

### 2.1. The INCOMMON mutation copy-number caller

We developed INCOMMON – *inference of copy number and mutation multiplicity for oncology* – a Bayesian model that can infer, for a tumour mutation, (i) the total copy number at the mutant locus, and (ii) the mutation multiplicity (i.e., the number of DNA copies that harbour the mutation). This information provides the mutant gene dosage and fraction, as canonically determined with copy number calling algorithms. INCOMMON takes input read counts data for *n* mutations *X* = {*x*_1_, …, *x*_*n*_} to develop the joint likelihood

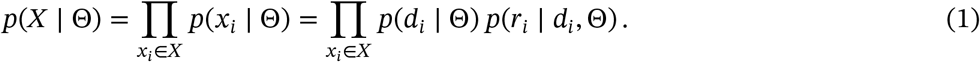

For every mutation *x*_*i*_ = ⟨*r*_*i*_, *d*_*i*_ ⟩, we use the number of reads *r*_*i*_ with the alternative allele and the total reads *d*_*i*_ (depth of sequencing). The model infers two sample-specific parameters, (*i*) the sample purity (0 < *π* ≤ 1), and (*ii*) the rate of reads per chromosome copy (*η* ∈ ℝ^+^), and two mutation-specific parameters, (*iii*) the tumour total copy number at the locus (*k*_*i*_ ∈ ℤ^+^) and (*iv*) the mutation multiplicty (*m*_*i*_ ∈ ℤ^+^, *m*_*i*_ ≤ *k*_*i*_).All mutations in a sample are modelled jointly, irrespective of their chromosomal location, borrowing information across multiple mutations to infer shared sample-level parameters and per-locus copy numbers more accurately. The model adopted by INCOMMON relates copy number to coverage linearly – the expected number of reads for *k* chromosome copies is *kη*-following the observation of a significant linear correlation (*R*^2^ ≥ 90% with p-value *P* < 0.05; Figure 1b) in 8,339 samples from the large-scale Pan-Cancer Analysis of Whole Genomes^27^ (PCAWG), Hartwig Medical Foundation^28^ (HMF) and The Cancer Genome Atlas^29^ (TCGA) cohorts, and following earlier RNA-based work^30–32^. INCOMMON links the number of reads to the multiplicity and total copy number (Figure 1c), considering tumour/ normal admixing

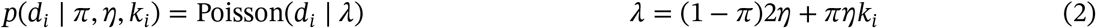

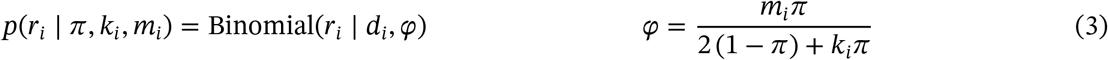

In this model, the sequencing depth follows a Poisson distribution with an expected number of reads *λ* defined by combining tumour and normal readouts. Given the depth, the number of mutant reads follows a Binomial distribution with success probability *φ* determined by the mixing of tumour and normal success rates. This construction induces overdispersion naturally through hierarchical modelling, avoiding the need for an arbitrary overdispersion parameter—a key advantage in terms of parsimony and interpretability. A graphical representation of the model, its priors and likelihood is shown in Supplementary Figure S1.

INCOMMON uses Markov Chain Monte Carlo sampling to estimate a posterior distribution over *m, k, η* and *π* (Figure 1d). To leverage the massive amount of public WGS data of human tumours and gain precision with targeted assays, we derived biologically informed prior distributions for (*k, m*) configurations from the PCAWG and HMF cohorts (examples for the most common alterations in Supplementary Figure S2, full distribution in Supplementary Table S1). For frequently mutated genes (e.g., *PIK*3*CA* and *TP*53 in breast cancer, *KRAS* in pancreatic cancer), these priors were gene- and tissue-specific (Supplementary Figure S3, Methods). Following a similar logic, the prior on *η* was derived empirically from the sequencing depth distribution of the analysed datasets (Supplementary Figure S4, Supplementary Table S2). Notably, we also modelled a prior for the input purity *π*, an essential parameter to estimate copy numbers. To support orthogonal data (e.g. from histopathological evaluation) but resist potential error in the input estimate, we centred a broad prior around the purity measurements provided with each sample (the 95% confidence interval lying within ±10-15 percentage points of the estimate). After posterior inference, INCOMMON uses posterior predictive checks to monitor the discrepancy between observed values and inferred posterior distributions on *k, m, η* and *π*. This information is used to annotate rare events (e.g. unexpectedly high copy numbers or multiplicities) and flag poor fits for the model before assembling all data in a detailed report of the analysis (Extended Data Figure 1). Examples of the report generated by INCOMMON MSK-MET samples, including some samples with a single detected mutation, are shown in Supplementary Figures S5, S6, S7 and S8, and discussed in detail in Supplementary Note 1.

We evaluated model performance using *N* = 50 iterations of random-split cross-validation (70% training, 30% test) across combined data from the PCAWG and HMF data (*n* = 6,492 samples, from 46 tumour types, Figure 1e and Supplementary Figure S9a). The model achieved a mean absolute error (MAE) of Δ*k* = 0.590 ± 0.016 for total copy number and Δ*m* = 0.201 ± 0.007 for mutation multiplicity, corresponding to deviations of less than one copy on average (i.e., one copy miscalled). In fact, 78.6% ± 0.9%of *k* predictions and 96.4% ± 0.4% of *m* predictions fell within ±1 copy of the true value, underscoring the model’s high predictive accuracy. The narrow variability across folds further highlighted the robustness and consistency of the estimates, supporting their reliability for downstream biological interpretation. We also compared INCOMMON calls with FACETS results for lower (LCN), major (MCN), and total copy number (TCN), which were publicly available for a subset of the whole MSK-MET cohort^33^ (Supplementary Figure S9b,c). While these statistics are lower-resolution than mutation multiplicities (unavailable in FACETS), we observed a good concordance from mean absolute deviations, which were 0.738, 1.00 and 1.94, respectively. Interestingly, concerning tumour purity, the two methods tended to adjust the pathologist’s estimate consistently, with a mean deviation of 9.72% between the two (Supplementary Figure S9d). INCOMMON demonstrated high computational efficiency (Supplementary Figure S9e), requiring a median of 42.0 ± 4.2 seconds and 90.607 ± 0.003 MB of RAM to analyse samples containing 10 mutations (median of 5 mutations per sample in MSK-MET). Runtime scaled linearly with input size, supporting its application in large-scale genomic pipelines (Supplementary Table S3).

### 2.2. The prognostic effect of gene mutant dosage across 20,000 human cancers

We used INCOMMON analysis from *n* = 21,937 tumours of the MSK-MET cohort to determine gene mutant dosage and its prognostic potential against overall survival (OS). INCOMMON jointly models copy number and mutational multiplicity, allowing the estimation of posterior probabilities for total and mutant allele configurations (*k, m*). Although this framework allowed survival analyses at full allelic resolution (Extended Data Figure 2, 3), stratification by individual values of *k* and *m* typically resulted in small and uneven subgroup sizes, limiting statistical power (average 36-37 for the most common diploid heterozygous and LOH configurations, less than 20 for others; Supplementary Figure S10a). Importantly, this constraint arises from sample size limitation rather than model design. To obtain a more robust approach and compare it with the canonical stratification into mutant (MUT) and wildtype (WT) tumours, we introduced a new statistic termed fraction of alleles carrying the mutation (FAM)

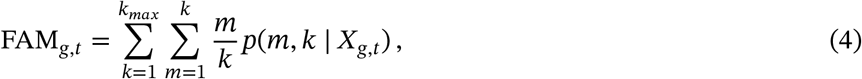

which is derived from the full posterior distribution of *p*(*m, k* | *X*_*g,t*_), computed by INCOMMON. This score, for a gene *g* and tumour type *t*, was paired with two cutoffs to determine high (FAM ≥ 75% for TSG, FAM ≥ 66% for oncogenes), low (FAM ≤ 25% for TSG and FAM ≤ 33% for oncogenes), and balanced dosage classes that maximise the rank statistics of the groups (Methods; Extended Data Figure 4). These groups are consistent with the canonical interpretation of allele-specific CNAs: the high dosage class includes, for example, deletions of the WT allele leading to loss of heterozygosity (LOH), typically occurring in TSGs^33–35^, and gains of the mutant allele frequently affecting oncogenes^33,34,36,37^, possibly combined as in copy-neutral LOH states; the balanced class includes mostly heterozygous diploid states, but also states of high ploidy with a similar number of mutant and WT alleles; low dosage reflects mutations following copy gains or post-mutation copy number events favoring the WT allele.

**Figure 3.**
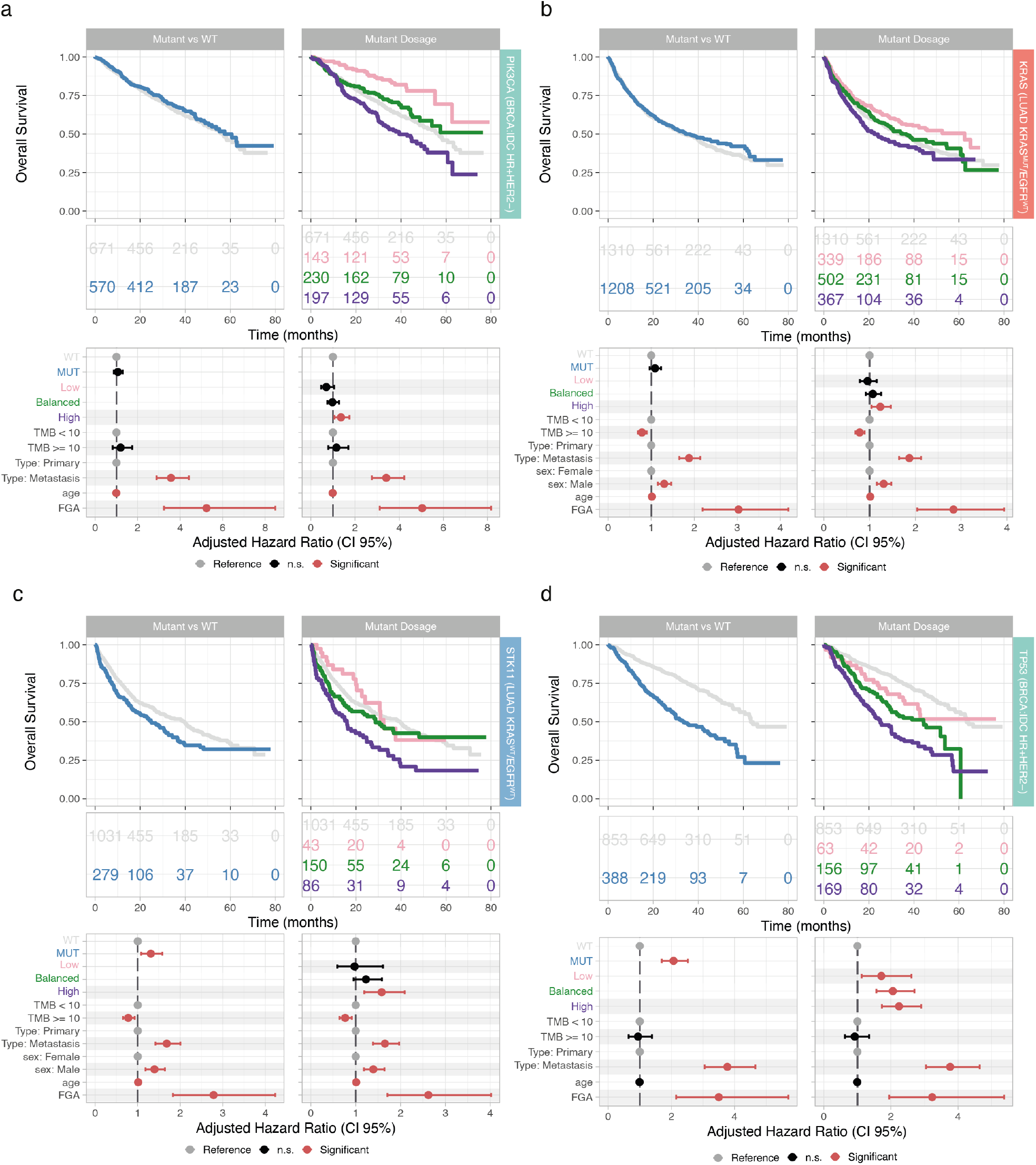
Gene mutant dosage refines survival analysis beyond mutation status. a. Comparison of Kaplan–Meier curves, annotated with numbers at risk, of overall survival (top panel) and adjusted hazard ratios (with 95% confidence interval, bottom panel) between mutant vs wild-type (left panel) and mutant dosage stratification (right panel), as in Figure 2e, for PIK3CA mutant patients with invasive ductal breast HR+HER2-carcinoma. For each patient group, the forest plot indicates whether the corresponding regression coefficient is significant (red) or not (black). The regression coefficient for the reference group (grey) is 1. b. Results for KRAS in EGFR wild-type lung adenocarcinoma. c. Results for STK11 in KRAS wild-type/EGFR wild-type lung adenocarcinoma. d. Results for TP53 in invasive ductal breast HR+HER2-carcinoma.

We first established the distribution of the maximum a posteriori values of read count per copy *η*, mutation copy number *k* and multiplicity *m*, FAM and sample purity across 286 genes and 50 tumour types in the MSK-MET dataset (Supplementary Figure S11). We then investigated the prognostic power of the classes derived from gene mutant dosage. Given the known different prognosis of cancer subtypes, we separately analysed those supported by sufficient sample numbers: mutant/wild-type *KRAS* and *EGFR* lung cancers, breast cancer molecular subtypes, micro-satellite stable (MSS) and hyper-mutant (HM) colorectal cancers, as well as endometrioid, serous and HM uterine cancers. A preliminary univariate analysis showed that the median OS was significantly different (false discovery rate, FDR, adjusted p-value *P* ≤ 0.1, Log-rank test) across mutant dosage classes in 16 cases. This covered 7 mutated genes (*APC, KRAS, NRAS, PIK*3*CA, RNF*43, *STK*11, *TP*53) regardless of the specific mutation position, and 2 hotspot mutations (*KRAS G*12 and *PIK*3*CA E*545*K*), for a total of 8 tumour types (Figure 2a). Interestingly, the lowest median OS was associated with the high dosage class in 75.0% of cases, and only rarely with the balanced and low classes (16.7% and 8.3%, respectively; Extended Data Figure 4; full analysis in Supplementary Table S4).

**Figure 2.**
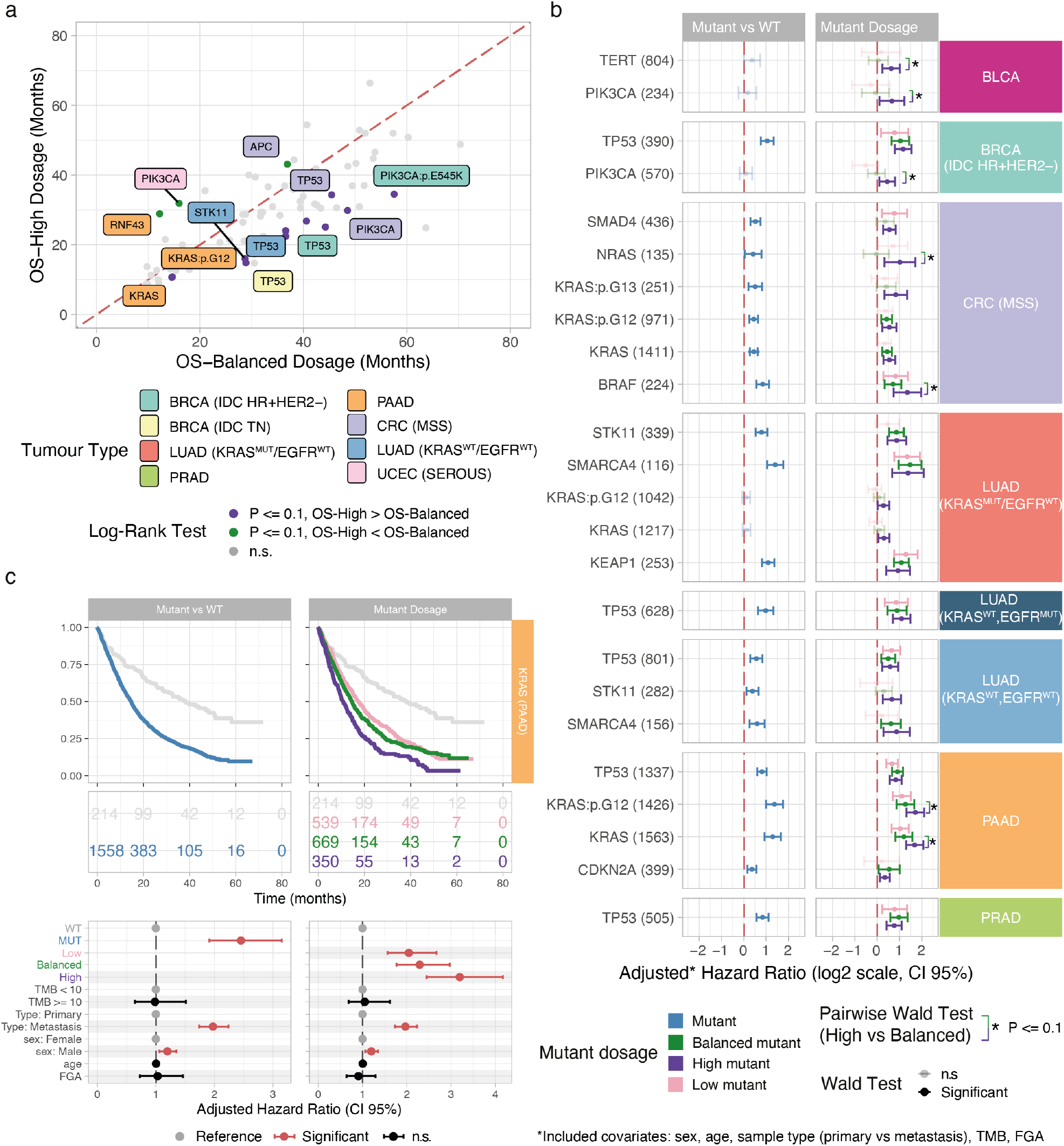
Prognostic effect of gene mutant dosage across 20,000 cancers. a. Comparison of median overall survival (OS) between high and balanced mutant dosage classes, highlighting markers with significant (Log-Rank test pairwise p-value *P* ≤ 0.1) OS difference. Tumour subtypes include hormone receptor-positive (HR+), HER2-negative (HER2-) and triple-negative (TN) breast cancer, lung adenocarcinoma (LUAD) with (MUT) or without (WT) KRAS and/or EGFR mutations, prostate adenocarcinoma (PRAD), pancreatic adenocarcinoma (PAAD), micro-satellite stable (MSS) colorectal cancer (CRC) and serous uterine corpus endometrial carcinoma (UCEC). b. Adjusted hazard ratios with 95% confidence interval (CI) from multivariate Cox regression accounting for sex, age, sample type (primary vs metastasis), tumour mutational burden (TMB) and fragment of genome altered (FGA), comparing canonical analysis of mutant vs wild-type tumours (left panel) with mutant dosage analysis (right panel). Markers with significant (two-sided Wald test, FDR-adjusted p-value *P* ≤ 0.1) negative prognostic impact associated with high mutant dosage are shown, highlighting those with a significant difference relative to the balanced dosage class. c. Comparison of Kaplan–Meier curves of OS (top panels) and adjusted hazard ratios between mutant vs WT (left panel) and mutant dosage stratification (right panel) for KRAS in pancreatic adenocarcinoma (PAAD).

Next, we tested this prognostic effect in a multivariable Cox model for OS adjusted for sex, age at diagnosis, sample type (primary versus metastasis), as well as two well-known biomarkers: the tumour mutation burden (TMB)^38–40^ and the overall aneuploidy^10,41–43^, measured as the fraction of genome altered (FGA). Since the association between gene dosage classes and survival may be confounded by higher TMB and FGA, adjusting for these two covariates was essential to understand the actual impact of gene mutant dosage statistics (Supplementary Figure S14).

For benchmarking, we compared mutant dosage against the purity-adjusted variant allele frequency (adjusted VAF), defined as the observed VAF divided by tumour purity – a simple approximation of allelic fraction used for patient stratification in targeted sequencing assays^44^. Mutant dosage consistently outperformed adjusted VAF in purity consistency, prognostic resolution, and precision (Supplementary Figure S10b-e, S12, examples in Supplementary Figure S13, Supplementary Note 2), Across tumour types, adjusted VAF identified 248 significant associations in univariate models and 121 in multivariate models (FDR < 0.1), compared to 260 and 147 for mutant dosage (+12 and +26 additional associations, respectively; Supplementary Table S10). Several biologically supported multivariate associations were uniquely recovered by mutant dosage, including high-dosage classes (*APC, BRAF*, S*MAD4* in CRC^45–47^; *EGFR* and *SMARCA4* in LUAD^48,49^; *CDKN2A* in PAAD^15^; *PTEN* and *ERBB3* in UCEC^50,51^, intermediate-dosage classes (*CDH1* and *GATA3* in BRCA^52,53^; *ARID1A* in PAAD and UCEC^15,54^; *FAT1* in CRC^55^), and low-dosage classes (*PIK3CA* in BRCA^56,57^; *TCF7L2* in CRC^58^; *TP53* in HCC and PAAD(^15,59^; *EGFR, KEAP1, SMARCA4* and *TBX3* in LUAD^44,48,49,60^). Beyond increased discovery power, the mutant dosage stratification provided by INCOMMON produced systematically narrower hazard-ratio confidence intervals, with a 16.93% smaller standard deviation of HR estimates on average. The largest differences occurred in high-dosage classes, where median standard errors in multivariate analysis were nearly twofold higher for adjusted VAF (0.579) than for mutant dosage (0.321, P < 0.0004). Simulations further showed that adjusted VAF is highly sensitive to tumour purity misspecification: as the simulated purity error increased, FAM estimation error rose to 50%, accumulating 10% additional error per 10% purity deviation, whereas INCOMMON remained comparatively stable (<20% error). Analytical derivations further showed that adjusted VAF and FAM coincide only under specific purity and copy-number configurations (Methods).

Our analysis showed that high mutant dosage was a negative prognostic factor in 24 cases (11 genes and 3 hotspots) across 8 tumour types (Figure 2b; full analysis in Supplementary Table S5). Among these, we searched for scenarios where the high dosage classes were significantly more associated with worse prognosis (higher adjusted hazard ratio, HR) compared to balanced ones. We found this strong trend for *KRAS* in pancreatic adenocarcinoma (*HR* = 3.19, *CI* = 2.45-4.17, *P* < 0.0001, Figure 2c), including G12 hotspot mutations (*HR* = 3.27, *CI* = 2.48-4.30, *P* < 0.0001), *BRAF* (*HR* = 2.55, *CI* = 1.67-3.90, *P* = 0.0001) and *NRAS* (*HR* = 2.02, *CI* = 1.25-3.27, *P* = 0.0144) in MSS colorectal cancer, *PIK*3*CA* (*HR* = 1.58, *CI* = 1.07-2.33, *P* = 0.0945) and *TERT* (*HR* = 1.55, *CI* = 1.18-2.03, *P* = 0.0140) in bladder cancer, and *PIK*3*CA* (*HR* = 1.36, *CI* = 1.07-1.74, *P* = 0.0325) in HR+HER2-invasive ductal breast cancer (Figure 3a). Notably, *PIK*3*CA* and *TERT* in bladder and breast cancers (Figure 3a) did not emerge as significant markers when using a canonical stratification of mutant versus wildtype patients, highlighting the enhanced resolution provided by gene mutant dosage.

A similar result occurred in specific lung cancer subtypes. For instance, high mutant *KRAS* dosage (*HR* = 1.23, *CI* = 1.04-1.46, *P* = 0.0384, Figure 3b) and its G12 hotspot mutations (*HR* = 1.22, *CI* = 1.02-1.46, *P* = 0.0328) were significanty prognostic in *EGFR*-wildtype adenocarcinomas, even if in this case we found only mild difference for the non-prognostic balanced class (*HR* = 1.06, *CI* = 0.899-1.26, *P* = 0.474).

The possibility of zooming into a mutant dosage analysis allows us to detect new prognostic classes which might be obscured in canonical analysis (mutant versus wildtype), increasing our biological understanding of tumour genomes. An exemplary case was that of *STK*11 in lung adenocarcinomas that are wildtype for the major oncogenes *KRAS* and *EGFR* (Figure 3c). In this case, we observed that only high mutant dosage *STK*11 was associated with negative prognosis (*HR* = 1.58, *CI* = 1.19-2.09, *P* = 0.0115). Similarly, in mutant *SMAD*4 colorectal MSS tumours, only high (*HR* = 1.46, *CI* = 1.20-1.78, *P* = 0.0006) and low (*HR* = 1.71, *CI* = 1.16-2.54, *P* = 0.0431) *SMAD*4 classes were prognostic, highlighting the strong impact of copy number alterations compared to plain *SMAD*4 mutations. Conversely, in mutant *KRAS* tumours – particularly those with G12 hotspot mutations – a low mutant dosage was not prognostic, unlike a balanced (*HR* = 1.36, *CI* = 1.17-1.58, *P* < 0.0001) or high dosage class (*HR* = 1.46, *CI* = 1.22-1.76, *P* = 0.0003), which were associated with an increased risk of death.

For the remaining markers where the high-dosage class was prognostic, we did not observe significant differences across dosage classes in univariate or multivariate analyses. The only exceptions were *TP*53 in certain breast, lung and prostate cancer subtypes (Figure 3d), where our mutant dosage stratification was significant in univariate analysis, but lacked robustness in multivariate analysis. In these particular cases, FGA and sample type accounted for most of the prognostic effect, suggesting at least strong collinearity between the presence of genome instability (high FGA) and *TP*53 mutations. In fact, we found that the FGA was progressively larger from low to balanced and up to high gene mutant dosage, the difference across dosage classes being significant (p-value *P* ≤ 0.1, Wilcoxon test) in HR+HER2- and triple-negative breast cancer, MSS colorectal cancer, hepatocellular carcinoma, *KRAS* wiltype lung adenocarcinoma, pancreatic and prostate cancer (Extended Data Figure 4).

### 2.3. Gene mutant dosage predicts metastatic propensity and tropism

We next examined how gene mutant dosage impacts metastatic propensity and organotropism. First, we compared the distribution of gene mutant dosage between primary (*n* = 13,216) and metastatic (*n* = 8, 721) tumour samples from the MSK-MET cohort.

Our analysis revealed an uneven distribution of INCOMMON gene mutant dosage classes between primary and metastatic tumours from 27 tumour types, 95 genes and 21 hotspot mutations (Figure 4), using a two-tailed Fisher’s exact test at significance level *α* = 0.1, together with a score *r* from the standardised residuals. Although the median enrichment was small for all classes (*r* = 0.732 for low mutant dosage in primary tumours, *r* = 0.136 for balanced dosage and *r* = 0.391 for high dosage in metastases), the distribution of scores was significantly different across classes (Extended Data Figure 5). As a general trend, low mutant dosage classes were more frequent in primary tumours, whereas high mutant dosage classes were enriched in the metastases (Supplementary Figure S15; full analysis in Supplementary Table S6).

**Figure 4.**
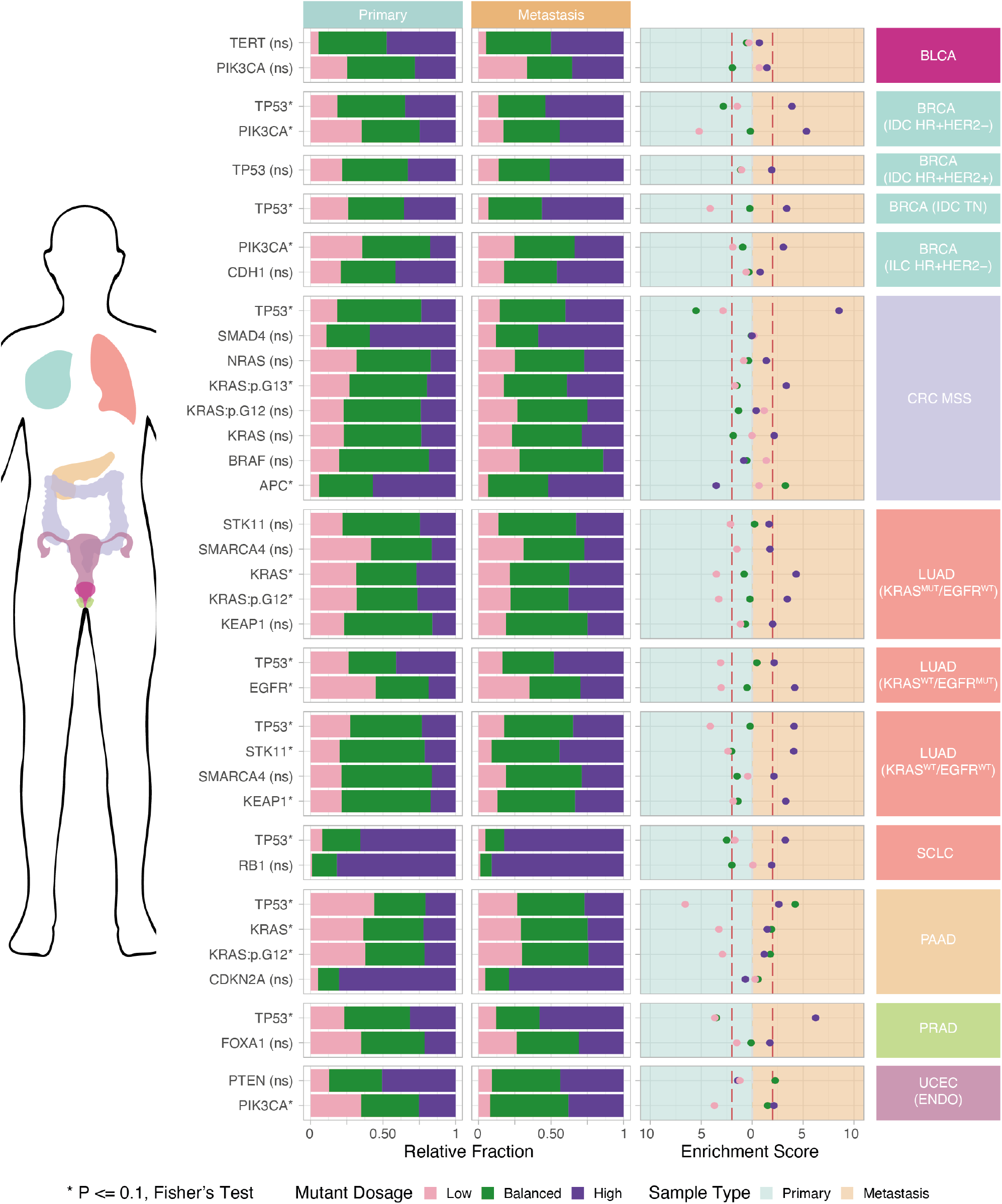
Enrichment of gene mutant dosage classes in primary tumour vs metastases. Distribution of low, balanced and high mutant dosage classes across 22 genes and 13 tumour subtypes, compared between primary and metastatic tumour samples (left panels). The right panel shows enrichment scores *r* for each class in either primary or metastatic tumours, evaluated through two-tailed Fisher’s exact test with significance level *α* = 0.1 and using standardised residuals. Vertical dashed lines indicate thresholds of large effect size (*r* = 2).

We observed a systematic enrichment of high mutant dosages in metastatic samples and low mutant dosages in primary samples, for both *TP53* and *PIK3CA* in breast and colorectal cancers, consistent with late biallelic inactivation of TSGs and amplified *PI3K* pathway activation, that might be associated with metastatic competence^26,28,61,62^. We found a similar enrichment pattern for *TP53* also in hepatocellular carcinomas, in pancreatic and prostate cancer, in melanoma, and in lung cancers together with *KRAS, EGFR* and *STK11*, all key regulators of *RAS*/MAPK or *RTK* signaling potentially implicated in metastatic dissemination^63^, and for *PIK3CA* in uterine endometrioid cancer along with *ARID1A*, a chromatin-remodeling gene in which biallelic alteration might be linked to progression^62^. Interestingly, in hypermutant uterine cancers, low dosages of *POLE* were enriched in primary samples and high dosages in the metastases, suggesting the potential role of *POLE* dosage in shaping tumour mutation rates^64^.

Additionally, high mutant dosage was enriched in several metastatic contexts: *APC* in prostate cancer metastases, indicating that WNT pathway deregulation might emerge as a later event^65^; *KEAP1* in lung cancer metastases lacking KRAS or EGFR mutations, suggesting that KEAP1 loss may be advantageous in metastatic niches^66^; *TP53* in small cell lung cancer metastases, that might be explained by the requirement for complete p53 pathway inactivation for late metastatic progression^67^; and KMT2A in melanoma metastases, where chromatin modification might play a central role in clonal diversification and metastatic fitness. Interestingly, in primary melanoma tumours, low dosages of *KMT2D, NTRK3, PREX2, ROS1, TP63* and *ZFHX3* were more frequent, a pattern compatible with these alterations acting earlier in tumour initiation rather than during dissemination^68^. Low dosages of *ZFHX3* were also enriched in primary prostate cancers^69^, and of *KRAS* in pancreatic cancers, potentially indicating that partial *KRAS* activation characterises early tumour development, with higher dosages emerging later in metastatic outgrowth^14–16^.

Overall, the strongest enrichment score in metastatic samples was for *TP53* in MSS colorectal (*r* = 8.52, *P* < 0.0001) and prostate cancers (*r* = 6.23, *P* < 0.0001), supporting prior observations that WT allele loss is necessary for proliferation toward metastasis^70^, and for *PIK3CA* in HR+HER2− breast cancers (*r* = 5.33, *P* < 0.0001), consistent with its role in chemotherapy resistance^71^. We validated our results on the MSK-MET cohort using *n* = 21,462 primary and *n* = 12,049 metastasis samples from the GENIE-DFCI cohort (Extended Data Figure 5, Supplementary Figure S16). A large fraction (89.29%) of the 56 significant markers that we found in MSK-MET had a consistent enrichment score between the two cohorts (inter-cohort variation lower than intra-cohort, Methods), confirming the general trend of low mutant dosage to be more frequent in primary tumours, and high mutant dosages in metastases across a total of *n* = 55,448 clinical samples.

To investigate the impact of gene mutant dosage on the metastatic propensity, we calculated the odds ratio (OR) of primary tumours with low, balanced or high mutant gene dosage to metastasise relative to wild-type (WT) tumours. In a preliminary univariate analysis, we found 12 genes and 4 specific hotspots with significant OR across 10 tumour types (Figure 5a). High mutant dosage increased the metastatic propensity in *n* = 9 cases (20.5% of the significant ones), and decreased it in *n* = 5 cases (Extended Data Figure 5). Low mutant dosage was primarily associated with reduced metastatic propensity (*n* = 11 cases), with only one case having a negative impact. The balanced class had, in most cases, a similar trend of the corresponding high or low counterpart, but with a less pronounced effect. To account for potential confounders, we applied a multivariable logistic regression model incorporating sex, age at sequencing, sample type, TMB and FGA, determining statistical significance using a Wald test with FDR-correction (Methods; Supplementary Figure S17; full analysis in Supplementary Table S7). The impact on metastatic propensity remained significant in 17 cases, including 8 genes and 4 hotspots across 8 tumour types (Figure 5b).

**Figure 5.**
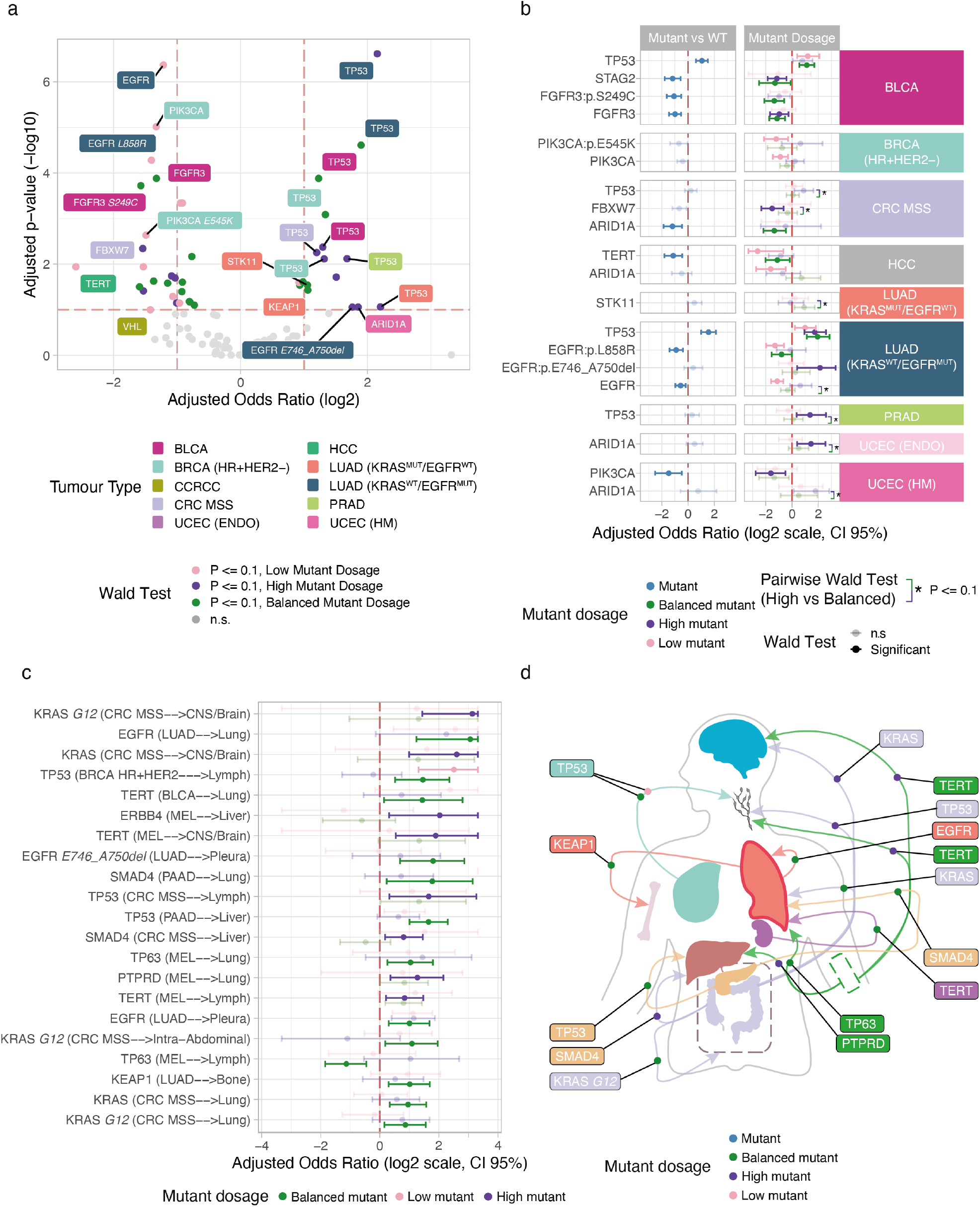
Gene mutant dosage predicts metastatic propensity and organotropism. a. Odds ratio (OR) of primary tumours with low (pink dot), high (purple dot) or balanced (green dot) mutant gene dosage, of giving rise to metastasis. The FDR-adjusted p-value (-log10 scale) against the OR (log2 scale) is reported. Lables highlighting genes and specific hotspots, with a significant impact on metastatic propensity, are colored according to the primary tumour type. b. Adjusted OR from multivariate logistic regression accounting for sex, age, tumour mutational burden (TMB) and fragment of genome altered (FGA), comparing canonical analysis of mutant vs wild-type tumours (left panel) with mutant dosage analysis (right panel). Markers with significant (Wald test, FDR-adjusted p-value *P* ≤ 0.1) impact on metastatic propensity associated with mutant dosage are shown, highlighting those with a significant difference between high and balanced dosage classes. c. Adjusted OR of metastatic tropism from primary tumours to other organs associated with mutant dosage. Significant cases are colored. d. Organotropic patterns associated with gene mutant dosage, represented by arrows starting from the primary site and pointing to the metastatic site. Dots are coloured according to the dosage class associated with the organotropic pattern, while labels highlight significant genes and specific hotspots, and are colored accordingly to the primary tumour type.

High mutant dosage was the only class significantly associated with higher metastatic propensity in several cases. In *KRAS*-wildtype lung adenocarcinomas, high-dosage *EGFR* mutations - particularly exon 19 deletions (*E746*_*A750del*) - were associated with a strong, albeit of borderline statistical significance, more than fourfold increase in metastatic propensity (adjusted odds ratio *OR* = 4.42, *CI* = 1.31 − 27.5, adjusted p-value *P* = 0.0884), suggesting that strong pathway activation might promote proliferation, stress adaptation, and invasive potential; by contrast, low-dosage mutations conferred reduced metastatic risk, suggesting that partial *EGFR* activation may suffice for tumor initiation but be insufficient to drive dissemination^72,73^. The effect was clear for *TP53* in prostate adenocarcinoma (*OR* = 2.59, *CI* = 1.27 − 6.00, *P* = 0.0450) and *ARID1A* in endometrial cancers (*OR* = 2.73, *CI* = 1.31 − 5.83, *P* = 0.0476), potentially representing double-hit events of these tumour suppressors that could enhance invasive potential^28,62^. Remarkably, these genes would not have emerged as markers of metastatic propensity if analysed without considering mutant dosage, i.e. by only comparing mutant vs WT tumours.

Consistent with results from the univariate analysis, several high-dosage mutant classes were associated with a trend toward reduced metastatic risk (*OR* < 1). In bladder cancer, high-dosage *FGFR3* (*OR* = 0.511, *CI* = 0.315 − 0.834, *P* = 0.0629) and *STAG2* (*OR* = 0.452, *CI* = 0.276 − 0.743, *P* = 0.0312) mutations halved the risk of metastasis, potentially identifying a category of tumours with luminal papillary phenotype and reduced cellular invasiveness^74–76^. Notably, high dosage *FBXW7* mutations in colorectal MSS cancers reduced metastatic risk by one third (*OR* = 0.350, *CI* = 0.199 − 0.644, *P* = 0.00734), as well as *PIK3CA* mutations in uterine cancer (*OR* = 0.327, *CI* = 0.146 − 0.711, *P* = 0.0381), supporting recent findings^77,78^.

More commonly, low dosage mutations were associated with reduced metastatic incidence. This was statistically significant for *TERT* (*OR* = 0.163, *CI* = 0.0392 − 0.635, *P* = 0.0111) and *ARID1A* mutations (*OR* = 0.324, *CI* = 0.151 − 0.716, *P* = 0.0111) in hepatocellular carcinoma, reflecting that partial loss of telomerase activity or chromatin remodelling may be insufficient to confer full metastatic fitness^62,79^. Low dosage *PIK3CA* mutations (*OR* = 0.541, *CI* = 0.363 − 0.806, *P* = 0.0177) including the *E545K* hotspot (*OR* = 0.432, *CI* = 0.226 − 0.811, *P* = 0.0340) in HR+HER2-breast cancers, similarly suggested that moderate pathway activation can support primary tumor growth but limit dissemination^71^. In *KRAS*-wildtype lung adenocarcinoma, low dosage *EGFR* mutations (*OR* = 0.457, *CI* = 0.329 − 0.636, *P* < 0.0001), including the *L858R* hotspot (*OR* = 0.406, *CI* = 0.254 − 0.654, *P* = 0.0004) were associated with decreased metastatic propensity, suggesting that partial *EGFR* signaling might suffice for tumor initiation but be insufficient to drive invasion and adaptation in distant niches^72^. The balanced dosage class showed reduced metastatic propensity in selected cases, including *FGFR3* (S249C mutations) in bladder cancer (*OR* = 0.392, *CI* = 0.232−0.661, *P* = 0.00271) and *ARID1A* in MSS colorectal cancer (*OR* = 0.392, *CI* = 0.222−0.726, *P* = 0.0308), indicating that an intermediate level of pathway perturbation may confer limited metastatic advantage^74–76^. Mutant TP53 tumours were generally more prone to metastasise than WT, irrespective of the mutant dosage, as in bladder cancer, *KRAS*-wildtype lung adenocarcinoma with *EGFR* mutations, and in MSS colorectal cancer^70^. In the latter case, while an increase in metastatic propensity for the high dosage relative to the balanced state was observed, its statistics remained below significance.

We then analysed organ-specific patterns of metastasis (Figure 5c,d), using multinomial regression adjusted for sex, age at sequencing, TMB and FGA (Methods; Supplementary Figure S18; full analysis in Supplementary Table S8). We found 33 patterns associated with mutant dosage, including 20 with strong statistical support (FDR-adjusted p-value *P* ≤ 0.05, Wald Test), and 13 showing borderline evidence (*P* ≤ 0.1), covering 14 genes and 4 hotspots across 5 primary tumours and 11 metastatic sites.

*CDH1* in HR+HER2-breast cancer displayed the strongest mutant dosage-linked tropism. Strikingly, both balanced (*OR* = 11.8, *CI* = 2.80 − 49.8, *P* = 0.00273) and high-dosage (*OR* = 9.58, *CI* = 2.09 − 43.8, *P* = 0.00837) classes conferred a roughly tenfold increase in ovarian metastasis, whereas low dosage showed no significant effect. This sharp contrast between dosages may suggest a threshold-like requirement for enhanced epithelial plasticity during peritoneal dissemination. In the same subtype, *GATA3* mutations showed a similarly strong but distinct pattern, with both balanced (*OR* = 3.24, *CI* = 1.32 − 7.98, *P* = 0.0311) and low dosage (*OR* = 10.4, *CI* = 3.06 − 35.7, *P* = 0.000331) predisposing to lung metastasis, indicating that intermediate activation may optimise migratory or lineage-plastic states required for pulmonary seeding. *ESR1* low-dosage mutations were protective against lymphatic dissemination (*OR* = 0.135, *CI* = 0.03 − 0.61, *P* = 0.0411), whereas *TP53* mutations showed a dosage-dependent increase in lymphatic tropism, with balanced (*OR* = 3.27, *CI* = 1.48 − 7.21, *P* = 0.0102) and low-dosage (*OR* = 7.56, *CI* = 2.26 − 25.29, *P* = 0.00185) classes associated with progressively higher odds of lymph-node metastasis. Together, these patterns suggested that partial impairment of *p53*-mediated differentiation programs may favour lymphatic seeding, whereas reduced perturbation of hormone-receptor may confer protection against lymphotropic spread^80^.

In MSS colorectal cancers, the odds of metastatic spread to central nervous system (CNS) metastases were more than fourfold for high dosage KRAS mutations (*OR* = 4.64, *CI* = 1.46 − 14.8, *P* = 0.0554), reaching a five-fold for G12 mutations (*OR* = 5.30, *CI* = 1.69 − 16.6, *P* = 0.00992), suggesting that KRAS-driven metabolic reprogramming and chromosomal instability might be linked to enhanced colonization of the CNS^81^.

Conversely, frequently harboured balanced *KRAS* mutant classes, which were also associated with a higher likelihood of intra-abdominal metastases, balanced *KRAS* dosage increased lung metastases (*OR* = 1.98, *CI* = 1.26 − 3.11, *P* = 0.0223), indicating dosage-specific division of metastatic routes, and that moderate pathway alteration may favour dissemination in the lung. *APC* mutations showed a different, largely suppressive pattern: balanced (*OR* = 0.353, *CI* = 0.19 − 0.65, *P* = 0.00412) and high-dosage (*OR* = 0.304, *CI* = 0.17 − 0.56, *P* = 0.000646) classes significantly reduced intra-abdominal metastasis, while high dosage tended to reduce ovarian dissemination (*OR* = 0.402, *CI* = 0.16 − 1, *P* = 0.0833). These effects, along with a borderline reduction in CNS metastases (*OR* = 0.270, *CI* = 0.08 − 0.87, *P* = 0.0694), suggested attenuated *WNT*-driven migratory potential at higher mutant dosages, with an inverse-dosage organotropism^81^. *PIK3CA* mutations displayed an opposite pattern: low dosage strongly increased ovarian metastasis (*OR* = 7.41, *CI* = 1.28 − 42.78, *P* = 0.0351), highlighting a low dosage optimum for *PI3K*-driven peritoneal spread^82^. In lung adenocarcinoma, multiple genes showed dosage-dependent behaviour. Balanced-dosage *KEAP1* mutations increased bone metastasis (*OR* = 2.22, *CI* = 1.3 − 3.8, *P* = 0.0117), whereas we observed a distinct pattern for high-dosage mutations, which increased head-and-neck metastases, albeit with borderline significance (*OR* = 6.26, *CI* = 1.12 − 35.0, *P* = 0.0737). This divergence highlighted a dosage-stratified organotropic spectrum, in which increasing levels of *KEAP1* inactivation may differentially support metastatic fitness across anatomical sites^83^. *EGFR* balanced-dosage mutations also increased bone metastasis (*OR* = 1.91, *CI* = 1.13 − 3.25, *P* = 0.0543), while high-dosage mutations increased liver involvement (*OR* = 2.03, *CI* = 1.1 − 3.74, *P* = 0.0574) and low dosage strongly increased mediastinal metastasis (*OR* = 3.94, *CI* = 1.29 − 12.09, *P* = 0.0327). The *E746*_A750del hotspot amplified these patterns, with high dosage predicting liver (*OR* = 5.04, *CI* = 1.62 − 15.65, *P* = 0.0103) and balanced dosage predicting pleural metastasis (*OR* = 2.52, *CI* = 1.09 −5.86, *P* = 0.0774). The *L858R* hotspot at balanced dosage further increased lung metastases (*OR* = 7.52, *CI* = 1.82 − 31.1, *P* = 0.0134). Together, these results revealed highly allele- and dosage-specific organotropic routes for *EGFR*-driven tumours. *KRAS* showed a complementary pattern: low-dosage mutations, whether general (*OR* = 0.328, *CI* = 0.15 − 0.70, *P* = 0.0104) or G12 (*OR* = 0.405, *CI* = 0.19 − 0.88, *P* = 0.0540), were associated with reduced odds of pleural metastasis, suggesting that intermediate levels of *RAS*-GTP signalling may be insufficient to support pleural colonisation. *PTPRT* and *PTPRD* mutations showed weaker but consistent effects: balanced-dosage *PTPRT* mutations were associated with reduced liver involvement (*OR* = 0.279, *CI* = 0.09 − 0.84, *P* = 0.0596), whereas high-dosage PTPRD mutations showed a borderline-significant increase in adrenal spread (*OR* = 3.90, *CI* = 1.15 − 13.24, *P* = 0.0576). Although modest in statistical strength, these patterns suggested that phosphatase-mediated modulation of signalling thresholds may contribute to organ-specific metastatic compatibility^84^. In pancreatic adenocarcinoma, both balanced (*OR* = 0.252, *CI* = 0.12 − 0.54, *P* = 0.00114) and low (*OR* = 0.439, *CI* = 0.21 − 0.91, *P* = 0.0802) *TP53* mutant dosage were associated with reduced intra-abdominal dissemination, and balanced dosage also corresponded to a lower likelihood of lymphnode involvement (*OR* = 0.216, *CI* = 0.08 − 0.56, *P* = 0.00262). Balanced-dosage *SMAD4* mutations showed strong lung tropism (*OR* = 4.30, *CI* = 1.48 − 12.53, *P* = 0.0375), suggesting that partial suppression of TGF-*β* signals might enhance epithelial-mesenchymal plasticity required for pulmonary seeding^85^. In melanoma, high (*OR* = 0.349, *CI* = 0.15 − 0.81, *P* = 0.0882) and balanced (*OR* = 0.212, *CI* = 0.08 − 0.55, *P* = 0.00917) *BRAF* mutations, including the V600E hotspot, were protective from liver metastases. *TP53* mutations showed an enrichment for CNS metastasis only at balanced dosage (*OR* = 0.208, *CI* = 0.07 − 0.65, *P* = 0.0399), indicating that modest *p53* impairment may facilitate neural-niche colonisation.

Overall, these findings demonstrate that metastatic tropism is not simply mutation-specific but is frequently dosage-dependent, with distinct dosage classes within a single gene often predisposing tumours to entirely different organotropic routes.

## 3. Discussion

In this study, we presented INCOMMON, a probabilistic framework to quantify gene mutant dosage across thousands of human cancers. Unlike existing copy-number callers^18–21^, which typically require access to raw sequencing data and are limited to matched tumour–normal assays, INCOMMON operates directly on variant-level read count data, thereby avoiding reliance on controlled-access sequencing files. Moreover, INCOMMON simultaneously infers mutation copy number and multiplicity, and uniquely incorporates informative prior distributions derived from large-scale whole-genome sequencing cohorts—a key innovation that enables robust and scalable inference across diverse datasets. By jointly modelling mutation copy number and multiplicity, our approach allows the measurement of gene mutant dosage, a statistic that provides a powerful and generalizable stratification of tumour genomes. We demonstrated the high accuracy of our method through extensive cross-validation using well-established WGS data from PCAWG and HMF, confirming the robustness of INCOMMON’s dosage assignments for downstream analyses. By applying this method to more than 50,000 samples from the MSK-MET and the GENIE-DFCI cohorts, we demonstrated that gene mutant dosage is a robust biomarker for patient outcome and tumour dissemination across multiple tumour types and genomic contexts.

Our analysis of over 20,000 tumour genomes from the MSK-MET cohort uncovered 46 tumour type-specific biomarkers for which dosage-derived classes were predictive of overall survival, including 38 with strong statistical support (FDR-adjusted p-value *P* ≤ 0.05, Wald Test), and 8 showing borderline evidence (0.05 < *P* ≤ 0.1). Notably, 13 of these associations were not detectable using conventional binary mutant/wild-type models. In most instances (70% of biomarkers), high mutant dosage was associated with significantly poorer prognosis, highlighting mutant dosage increase as a recurrent mechanism of aggressive tumour behaviour. This was particularly evident across the RAS/RAF pathway, where increased mutant dosage in *KRAS, NRAS*, and *BRAF* consistently tracked with adverse outcomes across multiple tumour types, extending and unifying prior observations from mutation- and CNA-based analyses^15,86–90^. These findings reinforce the emerging view that quantitative activation of RAS/RAF signalling—rather than the mere presence of a mutation—can be a key determinant of disease progression and may bear relevance for the use of RAS-targeted therapies^91^. Dosage effects also revealed context-dependent behaviour in genes such as *PIK3CA* and *TERT*. The prognostic impact of high *PIK3CA* dosage in HR+HER2− breast cancer aligned with recent results^56,57^, and with the tendency to increase metastatic risk. Conversely, in hypermutated uterine cancers, high dosage *PIK3CA* mutations were associated with a markedly reduced risk of metastasis and improved survival, possibly identifying a subset of endometrioid-like, estrogen-dependent tumours. In addition, the association of high dosage *PIK3CA* and *TERT* mutations with poor outcome in bladder cancer points to a broader relevance of dosage-mediated pathway activation in these tumours. That *TERT* dosage emerged as a prognostic marker is consistent with the unusually sensitive regulation of telomerase expression, where even modest dosage imbalances may meaningfully influence tumour adaptation^79^. Similarly, dosage-dependent effects in *STK11* clarified previously ambiguous prognostic patterns in lung adenocarcinoma^92^, revealing distinct interactions with oncogenic background that would remain obscured under standard mutant-versus-wild-type analyses. Finally, while *TP53* dosage showed prognostic associations in univariate analyses, multivariate models revealed that much of its apparent effect was absorbed by global aneuploidy measures. The observation that FGA progressively increases from low to balanced to high *TP53* dosage suggests a dosage-sensitive relationship between *TP53* disruption and genomic instability^93,94^, underscoring the need to account for background aneuploidy when evaluating the clinical impact of *TP53* alterations.

Altogether, these results extend prior findings that the number of mutations within single-copy regions or present in multiple copies provides stronger clinical predictive power than global measures of mutational burden or aneuploidy^95^. Importantly, they highlight how mutant dosage sharpens prognostic interpretation beyond coarse burden metrics^24,40^, though such metrics remain independent predictors in specific contexts^10^.

From the analysis of more than 50,000 unpaired primary and metastatic samples, we found that high mutant dosage was consistently enriched in metastatic tumours, most prominently for *TP53* and *PIK3CA* in breast and colorectal cancers. This enrichment, which we validated across the two independent MSK-MET and GENIE-DFCI datasets, was echoed by elevated metastatic odds ratios in multivariable models, supporting high dosage as a marker of metastatic fitness. For instance, *TP53* high-dosage mutations in prostate and MSS colorectal cancers^70,96^ showed the strongest enrichment in metastases and significantly increased metastatic propensity, even when controlling for global confounders like overall mutational burden and aneuploidy. Consistent with prior links between elevated *EGFR* dosage and advanced disease, we observed a clear dosage-dependent increase in metastatic potential, particularly for exon 19 deletions, while overall survival remained unchanged. This pattern illustrates how oncogene activation levels can fine-tune dissemination potential, creating a continuous phenotypic spectrum rather than a binary state. Although this diverges from earlier reports^72^, it is consistent with more recent studies showing no survival difference by *EGFR* dosage^73^. Our results suggest that survival outcomes in these tumours are instead primarily shaped by background aneuploidy and metastatic stage at diagnosis, which absorb much of the prognostic signal in multivariate models. The impact of *PIK3CA* mutant dosage on tumour behaviour appeared to be highly context-dependent. While high *PIK3CA* dosage was associated with both increased metastatic propensity and poorer survival in the HR+/HER2− breast cancers, in hypermutated uterine cancers, high-dosage *PIK3CA* mutations were linked to improved survival and a reduced risk of metastasis. These findings underscore the importance of tumour context, particularly mutation burden and immune environment, in shaping the phenotypic consequences of oncogene dosage. In uterine endometrioid cancer, high *ARID1A* mutant dosage was associated with increased metastatic propensity and showed a trend toward poorer overall survival, although it did not reach significance as an independent prognostic factor in multivariate analysis. This finding supports and extends prior observations that a double-hit mechanism—particularly loss of the wild-type *ARID1A* allele—may be required to drive malignant progression and promote metastasis^54,62,97^.

Beyond global metastatic risk, mutant dosage also predicted specific patterns of metastatic spread. We identified 33 tumour type–specific biomarkers, 20 with strong statistical support, for which mutant dosage stratified organotropic tendencies of metastasis, revealing a new layer of complexity in the molecular logic of organotropism and extending previous findings based on the independent analysis of mutations and copy number alterations^24^. In hormone-receptor–positive breast cancer, dosage-dependent alterations in epithelial adhesion, hormone signalling, and differentiation (across *CDH1, ESR1, GATA3, TP53*) channelled metastatic spread toward ovarian, pulmonary, or lymphatic routes, reflecting how graded perturbation of luminal-lineage circuitry shapes niche preference. In colorectal cancer, different dosage intensities across WNT, RAS/MAPK, and p53 pathways segregated tumours toward intra-abdominal, ovarian, pulmonary, lymphatic, or CNS dissemination, consistent with a modular architecture in which escalating pathway activation or suppression unlocks distinct metastatic competencies^81^. In lung adenocarcinoma, dosage variation across receptor tyrosine kinase and oxidative stress pathways (notably *EGFR, KEAP1, ARID1A, PTPRD, PTPRT*) delineated local thoracic versus distant organotropism, aligning with known metabolic and microenvironmental constraints of lung-derived metastases^44,83^. In melanoma, dosage-dependent tuning of MAPK, telomerase, and DNA-damage pathways coloured liver, lung, and brain tropism, suggesting that partial versus maximal pathway activation confers distinct microenvironmental compatibilities. In pancreatic cancer, dosage-driven modulation of *TP53* and *SMAD4* separated intra-abdominal, lymphatic, and pulmonary routes, highlighting how intermediate disruption of chromatin, EMT, or stress-response programs can redirect the metastatic trajectory.

These organotropic patterns—decipherable only through integrated mutation and copy-number analysis—open new avenues for understanding and potentially predicting the systemic spread of cancer.

## 4. Conclusion

Our findings support a model of oncogenesis in which the timing and allelic configuration of mutations—key drivers of mutant dosage changes—modulate tumour evolution, metastatic trait acquisition, and organ tropism. By highlighting gene mutant dosage as a clinically relevant axis of tumour genome interpretation, we demonstrate that not all mutations are equal: their dosage, shaped by clonal selection and copy number architecture, determines biological and clinical impact. Moving beyond binary mutation classification, our approach integrates mutation type with aneuploidy to yield a more precise risk stratification metric and a mechanistic explanation for previously unresolved clinical heterogeneity. Incorporating mutant dosage into diagnostic and prognostic models could enhance patient management, especially when treatment decisions depend on mutation status.

Despite the breadth of this new analysis, several aspects warrant further development to support broader clinical translation. Reference priors drawn from large WGS cohorts may need to be continuously refined to more closely reflect the histology, treatment exposure, and sequencing depth of the clinical populations under study. Inference of mutant dosage in tumours with pronounced intratumoral heterogeneity or very low purity remains challenging, even within our Bayesian framework, and represents an opportunity for methodological advancement. For example, the current model assumes that mutations and copy numbers are clonal (present in 100% of cancer cells). This approach is suboptimal for detecting ongoing evolutionary processes, which are better elucidated by subclonal deconvolution methods. Unfortunately, these signals are hard to extract from targeted panels and will have to be investigated in future extensions of our approach. In addition, variability in gene content and coverage across targeted sequencing panels currently limits the set of alterations for which dosage can be robustly inferred. The retrospective nature of the analysed cohorts also highlights the need for prospective validation of the prognostic and metastatic associations we uncovered. From a methodological perspective, moreover, extending the model to capture wildtype oncogene amplifications—though rare in primary tumours and not detectable in the present datasets—may provide additional insights into oncogenic pathway activation. The analysis of gene mutant dosage would also benefit from alternative assays, as transcriptomic and epigenomic read-outs may reveal whether high mutant dosage consistently translates into elevated oncogene expression or is tempered by compensatory mechanisms. Dissecting how high-dosage states modulate sensitivity or resistance to targeted agents, immunotherapies, and conventional chemotherapy should help turn dosage-based stratification into a practical precision-medicine tool. Matched longitudinal primary/metastatic samples would further refine our understanding of metastatic tropism, and survival data for the GENIE-DFCI cohort would enable validation of our MSK-MET-derived inferences.

In summary, this research advances key aspects of cancer genomics and presents INCOMMON as a transformative tool for interpreting targeted sequencing in the clinic. Our findings lay a strong foundation for future research and clinical translation, contributing both to personalised cancer therapy and to a deeper understanding of the biological complexity underlying genomic alterations. The clinical relevance of these insights underscores their potential to improve patient care.

## Data Availability

PCAWG and TCGA data used in this paper (mutations with matched validated CNAs) are available at https://doi.org/10.5281/zenodo.6410935, following 42.
MSK MetTropism data has been downloaded from cohort msk_met_2021 at the CBioPortal, following link https://www.cbioportal.org/study/summary?id=msk_met_2021.
AACR GENIE-DFCI data has been downloaded at https://www.synapse.org/# through access codes syn50678641, syn50678411, syn50678410, syn50678644, syn50678531, syn50678642, syn50678530, syn50678532, syn50678295, syn50678640, syn50678653, syn50678296.

## Data and software availability

All analyses presented in this paper, from data gathering to analysis results, are available in Zenodo at

10.5281/zenodo.20038793

PCAWG and TCGA data used in this article (mutations with validated matched CNAs) are available from Zenodo at

https://doi.org/10.5281/zenodo.6410935.

The MSK MetTropism data used in this article have been downloaded from the CBioPortal at

https://www.cbioportal.org,

under cohort `“msk_met_2021”` . AACR GENIE-DFCI data has been downloaded from Synapse at

https://www.synapse.org/

through access codes `syn50678641, syn50678411, syn50678410, syn50678644, syn50678531, syn50678642, syn5067853 syn50678532, syn50678295, syn50678640, syn50678653, syn50678296`.

HMF data (segmentation files and purity/ploidy estimates for *n* = 4,496 samples) were obtained from the Hartwig Medical Foundation (version 17/10/2020), using the request forms that can be found at

www.hartwigmedicalfoundation.nl

The files were generated using the PURPLE pipeline, available at

github.com/hartwigmedical/hmftools/

We then manually converted the Hartwig annotations to match those in ICGC (Supplementary Table S9).

INCOMMON is an open-source R package downloadable from Github at

https://caravagnalab.github.io/INCOMMON

The tool webpage contains RMarkdown vignettes to run analyses, visualise inputs and outputs, and parametrise the tool.

INCOMMON is also available as a ShinyApp that can be used to browse analysis results and run online similar analyses. The app is available online at

https://ncalonaci.shinyapps.io/incommon/

## Author contributions

NC and GC conceptualised and formalised INCOMMON, which was implemented by NC with support from DC, SM and SS. NC, BR, SS, MMS, AS, TG and KB gathered real data, which was analysed by NC, EK, DC, GG, GC and LE, and interpreted by all authors. GC supervised the project. NC, EK and GC drafted the manuscript that all authors approved in final form.

## Funding and acknowledgments

GC discloses support for the research of this work from the Associazione Italiana per la Ricerca contro il Cancro (AIRC) [MFAG number 24913 and Bridge number 32107]. EK discloses financial support for this work through funding from the institutional “Ricerca Corrente” granted by the Italian Ministry of Health. MM-S discloses support for the research of this work from the Associazione Italiana per la Ricerca contro il Cancro (AIRC) [MFAG number 22940 and IG number 32059], and from the Italian Ministry of Health [PNRR-MCNT2-2023 number 12377963]. KB discloses support for the research of this work from the Swiss National Science Foundation [P500PM number 217647/1]. AS discloses support for the research of this work from the Associazione Italiana per la Ricerca contro il Cancro (AIRC) [IG number 28961] and by the European Research Council (ERC) [Consolidator Grant number 101125077]. TG discloses support for the research of this work from Cancer Research UK [DRCNPG-May21 number 100001]. NC, DC, SM, GG, SS, LE and BR declare no relevant funding. This publication and the underlying study have been made possible partly based on data that Hartwig Medical Foundation has made available to the study through the Hartwig Medical Database. We thank the reviewers and editor for providing constructive feedback to improve our manuscript.

## Inclusion and ethics

All collaborators of this study who have fulfilled the criteria for authorship required by Nature Portfolio journals have been included as authors, as their participation was essential for the design and implementation of the study. Roles and responsibilities were agreed among collaborators ahead of the research.

## 5. Methods

### Datasets Used for Prior Construction and Performance Evaluation

To compute the empirical prior distributions of copy number configurations (*k, m*), we assembled a cohort of high resolution whole-genome sequencing data, collecting samples from the Pan-Cancer Analysis of Whole Genomes^27^ dataset (PCAWG, *N* = 2,777 samples, 40 tumour types, median depth 45 and purity 65%), and from the Hartwig Medical Foundation^26^ (HMF, *N* = 8,616 samples, 28 tumour types, median depth 96 and purity 55%) dataset. We retrospectively assembled CNA calls from PCAWG, which were previously obtained as part of a curated consensus(Dentro et al. 2021), and CNA calls from HMF that were previously obtained using PURPLE^26^. We used the resulting dataset of mutations and CNAs as the benchmark dataset *B*. Importantly, this aggregated dataset includes samples from both primary (from PCAWG) and metastatic tumours (from HMF), ensuring its usefulness in the analysis of datasets containing both sample types, such as MSK-MET. Additionally, we used data from The Cancer Genome Atlas^29^ (TCGA, *N* = 1,068 samples) as a validation dataset. As the method is fully data-driven, these empirical priors can be readily updated or refined as additional high-quality datasets become available.

### Bayesian inference of copy number and mutation multiplicity using INCOMMON

INCOMMON is a Bayesian framework to infer mutation multiplicity and copy number from read count data. The data consist of a set of *n* mutations *X* = {*x*_1_, …, *x*_*n*_} from a tumour sample, with each mutation *x*_*i*_ = ⟨*r*_*i*_, *d*_*i*_ ⟩ characterised by the number of reads with the variant *r*_*i*_ ∈ ℤ^+^ and the total number of reads *d*_*i*_ ∈ ℤ^+^ (sequencing depth). INCOMMON jointly models the purity 0 < *π* ≤ 1 and the rate of reads per chromosome copy (*η* ∈ ℝ^+^) of the sample, and the total copy number (*k*_*i*_ ∈ ℤ^+^) and mutation multiplicity (*m*_*i*_ ∈ ℤ^+^, *m*_*i*_ ≤ *k*_*i*_) of each mutation in the sample. The joint likelihood of the model is

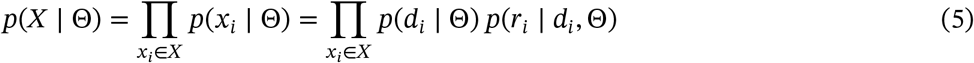

where Θ is the vector of model parameters Θ = (*π, η*, {*k, m*}^*n*^).

In INCOMMON, the total number of reads follows a Poisson distribution

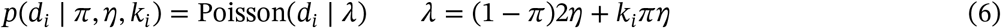

The expected value *λ* is contributed by reads from the normal cells, present in the sample in a fraction 1 − *π*, and from tumour cells present in a fraction *π*. The expected number of reads from normal cells, assumed to be diploid, is 2*η* across the whole genome. The expected number of reads from tumour cells is *kη*, assumed to be proportional to the unknown total copy number.

Given the total read count *d*_*i*_, the number of reads with the variant *r*_*i*_ follows a Binomial distribution

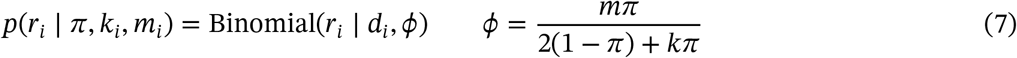

The probability of collecting a read with the variant (success probability) *ϕ* is equal to the variant allele frequency (VAF) expected for a mutation with copy number configuration (*k, m*), adjusted for the sample purity. INCOMMON embeds prior knowledge of the model parameters Θ using a prior distribution that can be factorised into

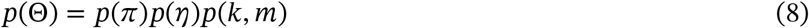

INCOMMON uses Markov Chain Monte Carlo (MCMC) sampling to estimate the model posterior *p*(Θ ∣ *X*), that would otherwise be intractable analytically.

### Empirical Priors

#### Copy number and mutation multiplicity

A key feature of INCOMMON is the use of biologically informed prior distributions for copy number and mutation multiplicity configurations across genes and tumour types. These priors were derived from a training dataset of 4543 samples (95896 mutations with available copy number states) curated from two large high-resolution whole-genome sequencing cohorts: the Pan-Cancer Analysis of Whole Genomes (PCAWG, 1,519 samples) and the Hartwig Medical Foundation (HMF, 3,024 samples). A subset of samples was held out for model performance evaluation.

We empirically estimated gene- and tumour type-specific prior distributions *p*(*k, m*) over 36 possible combinations of total copy number and mutation multiplicity, with an upper bound of *k*_max_ = 8. The prior probabilities were computed based on the observed frequencies of each configuration in the training dataset for each tumour type. To ensure statistical robustness, we required that a gene be mutated in at least 50 samples within a given tumour type to construct a tumour-specific prior; otherwise, a pan-cancer prior was used by aggregating data across all tumour types.

Missing observations were imputed using pseudocounts, estimated through a Gaussian-kernel smoothing approach that incorporated copy number and mutation multiplicity distances. In particular, given observed mutation counts *n*_*i*_ for different (*k, m*) configurations, we defined a custom distance metric 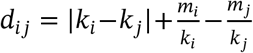 and applied a Gaussian kernel 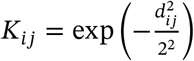 to weigh the observations. We then computed the pseudocounts as 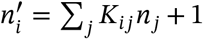 where we used 1 as the base pseudocount, ensuring numerical stability. To preserve the overall mutation burden, we applied a normalisation step to get the final pseudo-count 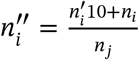.

#### Tumour purity

The inference of copy number and multiplicity with INCOMMON critically relies on the sample purity, a piece of information that is usually accessible as provided together with genomic data. However, the available purity estimates are often pathology-based and, thus, potentially incorrect or differing from molecular estimates. The full Bayesian formulation of INCOMMON allows using an informative, yet broad, prior on sample purity *π*, defined starting from the available estimate *π*_0_. INCOMMON uses a Beta distribution over purity

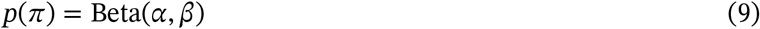

Here, for each sample, we choose specific values of the shape parameters *απ* such that the mean of the distribution is equal to the available estimate *π*_0_, and the expected variance is fixed to *σ*^2^, namely:

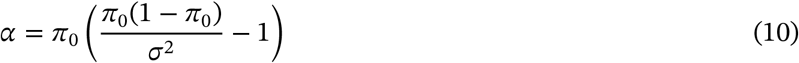

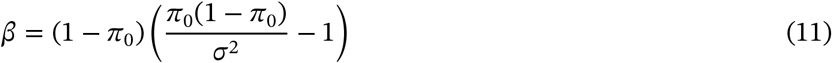

We used *σ*^2^ = 0.05 for all the datasets we analysed. In this way, INCOMMON accounts for uncertainty in purity estimation of around 10%. Potentially, there can be cases in which the input purity estimate is not supported by the inference of INCOMMON, despite the allowed uncertainty. We control for these extreme cases through posterior predictive checks (see, for an example, Extended Data Figure 1).

### Read count rate per chromosome copy

For the read count rate per chromosome copy *η*, we estimate the prior distribution from the data in an Empirical Bayes fashion. In principle, following Equation (**??**) the expected total number of reads from a mutation site with copy number *k*^′^ is *E*(*d*_*i*_ ∣ *k* = *k*^′^) = 2(1 − *π*) + *k*^′^*π η*. Thus, an empirical estimator of *η* of the per-copy rate can be obtained from the mean total number of reads coming from all the mutations with *k* = *k*^′^ as 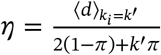. Since a priori we do not know the copy number of mutations in the sample, the best we can do is to use the expected value with respect to the prior probability 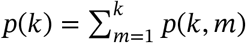.

In this way we obtain the estimator

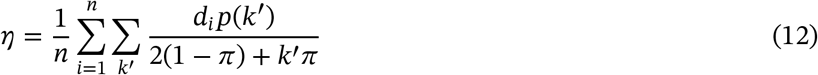

We then use a Gamma prior distribution over *η*

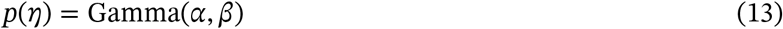

in which we choose the values of the shape parameters such that the mean is equal to our empirical estimation 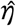 and the variance is equal to the empirical variance Var(*η*) across the dataset

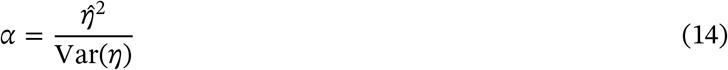

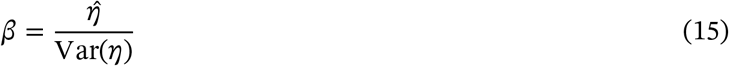

This prior reflects our prior knowledge over latent copy number and multiplicity configurations *p*(*k, m*) and adapts it to the observed distribution of sequencing depth in the dataset, constraining posterior estimates of *η* to reasonable values. The prior distribution over *η* for all the analysed cohorts (MSK-MET, DFCI) are reported in Supplementary Figure S4, along with the corresponding observed distributions of sequencing depth. The use of an informative prior for the estimation of the per-copy count rate is crucial, especially in settings where normal samples are not available (due to questions of data disclosure or in tumour-only assays) so that depth ratios cannot be computed.

### The fraction of alleles with the mutation (FAM)

INCOMMON allows for the estimation of mutation copy number and multiplicity. In our study, utilising INCOMMON, we estimated the Fraction of Alleles carrying the Mutation (FAM), representing the relative fraction of mutant alleles of a gene within all cells of the sample. The probabilistic model enables the calculation of the expected value of FAM as:

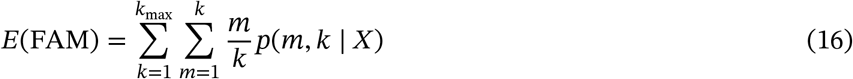

where *p*(*m, k* ∣ *X*) denotes the joint posterior probability of *k* and *m*, obtained by marginalizing the full posterior over sample purity *π* and read count rate per chromosome copy *η*,

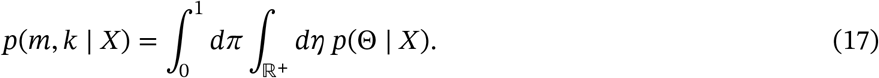

Even in cases where the exact configuration of (*k, m*) cannot be precisely determined, the estimator in Equation (16) leverages the full posterior distribution to provide a robust estimate of their ratio *m*/*k*, weighting contributions from different configurations according to their joint probability.

### Classification based on FAM cutoffs

To determine the optimal thresholds for dichotomising FAM based on overall survival (OS), we used the maximally selected rank statistics method. We searched for the lower cutoff in the range FAM < 0.5 and for the upper one in the range FAM > 0.5. The maximally selected rank statistics method identifies the cutpoint that yields the most significant split in survival, while correcting for multiple testing. We required at least 20% of all samples in each group, and adjusted models for age, sex, tumour mutational burden, fraction of genome altered, and sample type (primary tumours vs metastases). Analyses were conducted using R version 4.3.2 with the tidyverse (v2.0.0) and survminer (v0.4.9) packages.

### Benchmarking against adjusted VAF

To benchmark mutant dosage against the conventional tumour purity-adjusted VAF (VAF/*π*) stratification, we compared both approaches in terms of purity robustness and prognostic resolution. Adjusted VAF values were often inconsistent with purity estimates (189,656 mutations with VAF ≤ *π* vs 34,684 with VAF > *π*), whereas INCOMMON jointly infers purity and allele configuration, reducing such implausible cases to 17,404 out of 206,936 (Supplementary Figure S11a). When rank-based cutoffs were optimized for adjusted VAF, the dynamic ranges obtained (0.36–1.33 for TSGs and 0.24–1.35 for oncogenes; Supplementary Figure S11b) were comparable, but the number of significant prognostic classes was lower than for mutant dosage (+12 and +26 additional classes, respectively; Supplementary Figure S11c). Moreover, Cox regression analyses revealed wider 95% confidence intervals for hazard ratios with adjusted VAF, indicating reduced precision (Supplementary Figure S11d). Representative cases, including one in which adjusted VAF outperformed mutant dosage, are shown in Supplementary Figure S12.

Although purity-adjusted VAF may resemble the FAM score in specific cases, the two quantities are analytically distinct. The expected VAF, conditioned on tumor purity *π*, mutation multiplicity *m*, and total copy number *k*, is

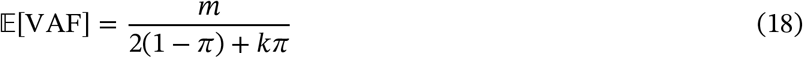

The adjusted VAF equals the FAM only under restricted purity and copy-number configurations. Taking limits over tumor purity yields

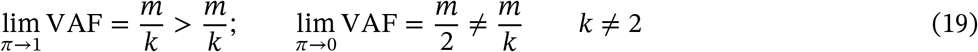

Thus, at high purities the adjusted VAF systematically overestimates the fraction of mutant alleles, whereas at low purities it converges to approximately half the FAM value for *k* = 1 and diverges for other copy-number states. This demonstrates that the adjusted VAF and FAM coincide only approximately and that the latter provides a more robust and biologically interpretable measure of mutant dosage.

### Posterior Predictive Checking and Bayesian p-value Calculation

To assess model fit and evaluate potential discrepancies between observed data and posterior predictive distributions, we performed posterior predictive checks and computed Bayesian p-values.

#### Posterior Predictive p-value

For each mutation-specific observed quantity *y*_*i*_, such as reads with the variant *r*_*i*_ or total reads *d*_*i*_, INCOMMON generates *S* posterior predictive samples 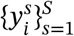 from the model posterior distribution, and evaluates the posterior predictive p-value as

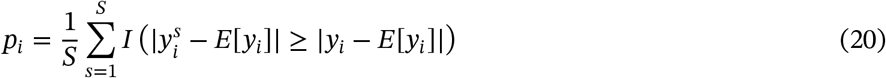

where 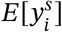 is the posterior mean of the replicated observations. The indicator function *I*(⋅) evaluates to 1 if the absolute deviation of the posterior predictive sample from the posterior mean is at least as large as the deviation of the observed data and 0 otherwise. This p-value thus quantifies the extremity of the observed data under the model posterior, providing a measure of the goodness of fit for individual mutations.

In samples with only one or two observed mutations, posterior predictive p-values for the Poisson depth model may approach 1, as the inferred copy number *k* closely reproduces the single observed depth; this reflects the limited data rather than model misfit. INCOMMON remains applicable to such cases: the Bayesian formulation provides natural regularization, as the sample-level parameters (purity and per-copy read rate) are informed by their priors, yielding stable and biologically plausible posteriors that appropriately reflect increased uncertainty. Examples of single-mutation cases are shown in Supplementary Figures S7–S8.

#### Bayesian p-value

At the level of global (sample-wise) parameters *y*, such as tumour purity *π* and read counts per chromosome copy *η*, INCOMMON estimates Bayesian p-values by comparing posterior and prior distributions. Given *S* posterior samples 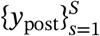 and prior samples 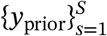, the test statistic is based on deviations from the prior mean *E*[*y*_prior_].

INCOMMON computes the test statistic differently depending on the assumed prior distribution:

For a Beta prior:

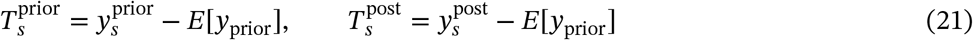

For a Gamma prior:

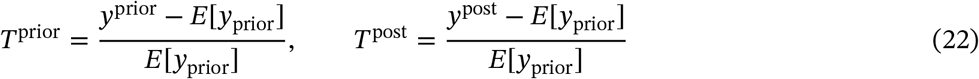

The Bayesian p-value is thus computed as the proportion of posterior test statistics that exceed or equal the prior test statistics:

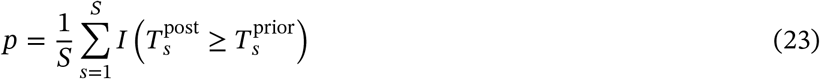

This Bayesian p-value quantifies the extent to which posterior samples deviate from prior expectations, providing an additional measure of model adequacy.

### Evaluation of model performance

We developed an iterative approach to evaluate the performance of INCOMMON and the impact of empirical priors on the model predictions. Our method involved iteratively splitting the benchmark dataset B into two subsets: the prior dataset P (70% of the samples) for prior estimation, and the evaluation dataset E (30% of the samples) for performance evaluation. By aggregating results over *N* = 50 iterations, we computed the mean absolute error (MAE) of INCOMMON predictions of mutation total copy number and multiplicity along with their standard deviations.

### Setting an upper bound for total copy number *k*_*max*_

In INCOMMON, the model likelihood is computed over a discrete set of combinations of total copy number *k* and multiplicity *m*. While for multiplicity, an upper bound is set naturally by the constraint that *m* ≤ *k*, i.e. the number of mutant copies cannot exceed the total number of copies, for the total copy number, the choice of the upper bound *k*_*max*_ is arbitrary. In order to choose a reasonable value of *k*_*max*_, we used the benchmark dataset B to evaluate the frequency of large (> 4) values of *k* and set a threshold above which the number of mutations is negligible. As shown in Supplementary Figure S19, the distribution of total copy number values significantly varied between wild-type (WT) and mutant sites, with the number of sites decreasing more rapidly with *k* for the mutant sites with respect to WT. In particular, we found fewer than *n* = 100 mutations per value of *k* for *k* > 8 across the whole benchmark dataset (*N* = 2,375,815 mutations in total). We thus set the upper bound to *k*_*max*_ = 8 for all our analyses.

### Survival Analysis

Overall survival (OS) time (months) and status (living vs dead) for the MSK-MET were taken from the publicly available clinical release. OS curves and OS values were calculated using the univariate Kaplan–Meier estimator. All p-values and adjusted hazard ratios were evaluated through multivariable Cox proportional hazards models accounting for covariates including age at sequencing, sex, tumour mutational burden per megabase (TMB) and fragment of genome altered (FGA). Multivariable Cox proportional hazards models were constructed for each gene, and p-values of mutant dosage classes (WT, MUT, Low, Balanced, High) and all covariates were adjusted by FDR independently for each tumour type using the number of genes that were analysed simultaneously. For each tumour type, all genes that were mutated in less than 150 samples were excluded from the analysis. The same threshold was used for specific recurrent amino acid substitutions (hotspot mutations). Analyses were conducted using R version with the tidyverse (v2.0.0), survival (v3.7-0), survminer (v0.4.9) and multcomp (v1.4-26).

### Enrichment in primary tumours vs metastases

Sample type (primary tumour/metastasis) annotations for the MSK-MET were taken from the publicly available clinical release. To evaluate whether any of the three gene mutant dosage classes were enriched in either primary tumours or metastases, we performed an exact Fisher’s test. For each gene and tumour type, we constructed a two-way contingency table of sample counts between class and sample type, and applied Fisher’s test to assess deviation from independence under the null hypothesis. We evaluated an enrichment score from the standardised residuals obtained from the contingency table, which measures the deviation of observed from expected counts, normalised by the standard error. P-values for all the tests were FDR-adjusted through the Benjamini-Hochberg method, independently for each tumour type, using the number of genes that were analysed simultaneously. Analyses were conducted using R version 4.3.2 with the tidyverse (v2.0.0) and stats (v4.3.3).

### Metastatic propensity and organotropism

Patient status (metastatic/non-metastatic) and site of metastasis annotations for the MSK-MET were taken from the publicly available clinical release. To assess associations between gene mutant dosage class and sample type (metastatic propensity), univariable and multivariable logistic regression models. The univariable model included only gene mutant dosage as a predictor. The multivariable model included covariates for age at sequencing, sex, tumour mutational burden per megabase (TMB) and fragment of genome altered (FGA). All models were specified as generalised linear models using the binomial family with a logit link function. To evaluate associations between gene-specific mutant dosage and patient status (organotropism), we used a multinomial log-linear model with metastatic site as the (multivariate) outcome variable and mutant dosage (multivariable) as the predictor. For each gene, the most frequent metastatic site in the wild-type class was used as the reference category. From the fits, we extracted coefficient estimates for all variables and 95% confidence intervals. A two-sided Wald test for statistical significance was performed for each regression coefficient. P-values for all the covariates were FDR-adjusted through the Benjamini-Hochberg method, independently for each tumour type, using the number of genes that were analysed simultaneously. Wald pairwise comparisons between mutant dosage classes were performed using general linear hypothesis testing. Analyses were conducted using R version 4.3.2 with the tidyverse (v2.0.0), stats (v4.3.3), nnet (v7.3-20) and multcomp (v1.4-26).

### Exploratory heuristic for identifying potentially subclonal mutations

Since INCOMMON assumes mutations to be clonal (cancer cell fraction CCF = 1), we explored a simple post-hoc heuristic to assess whether its posterior predictive checks (PPCs) could flag potentially subclonal variants. Specifically, we considered mutations that failed the PPC for variant-supporting reads and whose observed counts were lower than predicted after accounting for copy number and multiplicity. To minimize confounding, we restricted this set to mutations with maximum a posteriori multiplicity m=1 and to samples that passed the PPC for purity, ensuring that apparent deficits could not be explained by inflated multiplicity or inaccurate purity inputs. Eventually, the number of potentially subclonal events per gene-tumour-type combination was extremely small – typically 1-10 mutations, and generally <5% of all mutations in the group. Due to such limited sample sizes, and the inability to confidently separate true subclonality from residual technical noise, we concluded that INCOMMON outputs do not currently support clonality-stratified survival and metastatic tropism analyses. Systematic estimation of CCF is therefore reserved for future work.

## 6. Extended Data Figures

**Extended Data Figure 1.**
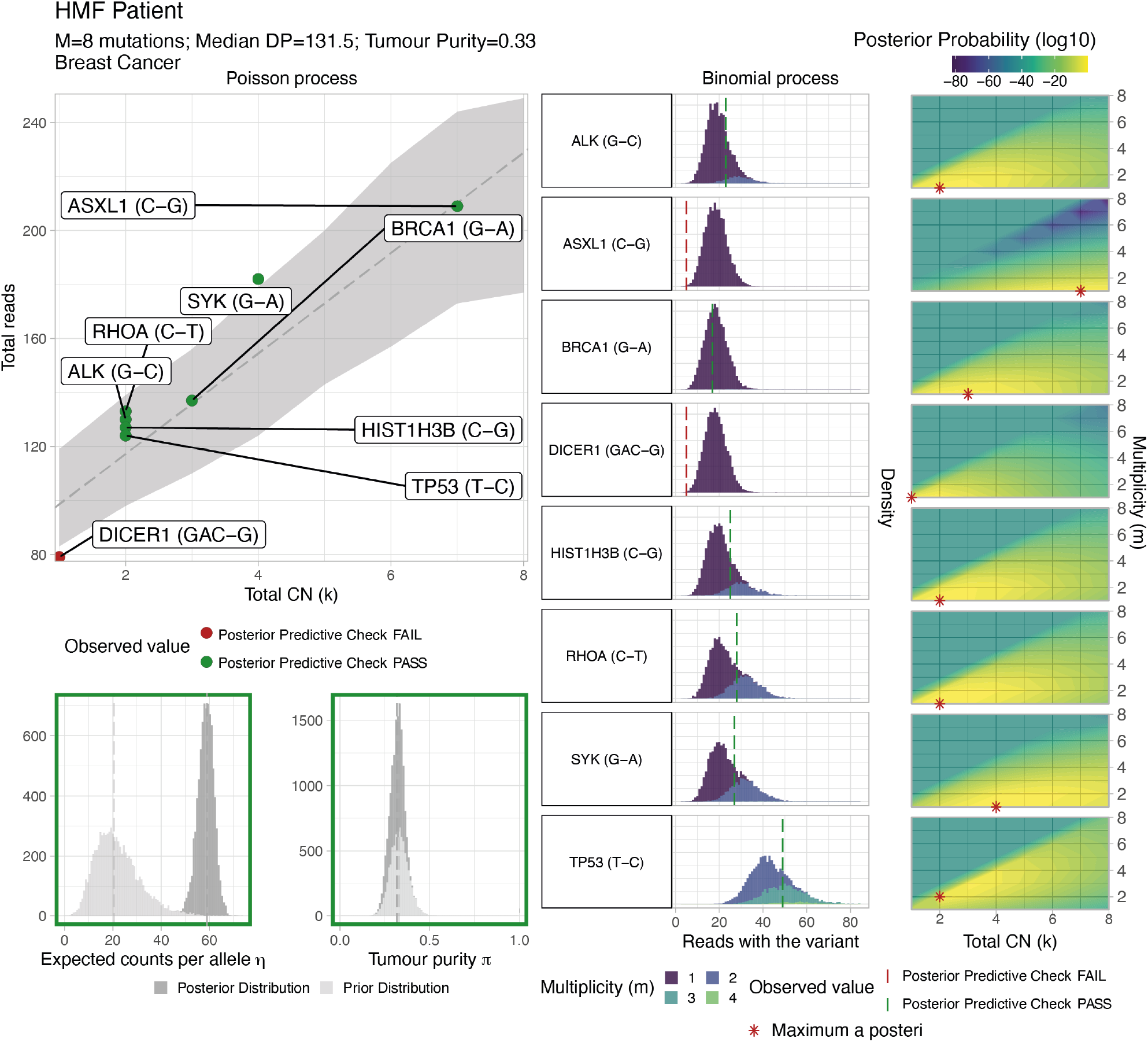
Report of INCOMMON inference for a tumour. Report of the INCOMMON fit of a breast cancer (BRCA) sample from HMF, containing 8 mutations sequenced at medain depth 131.5 and with pathologist’s tumour purity estimate of 0.33. In the top left panel, the report displays the Poisson likelihood of total read counts parametrised by mutations’ total copy number. The grey line and ribbon represent the expected number of reads per *k* chromosome copies-*kη*, and the 95% confidence interval of the posterior distribution of total reads per value of *k*. Red dots highlight data failing the posterior predictive check (PPC). The central panel displays the posterior distribution of reads with the variant for all sampled values of multiplicity *m*, with the vertical line indicating the observed value, in red if failing the PPC. The bottom left panels show the comparison between the prior and posterior distributions for the expected count per allele *η* and the tumour purity *π*. The rightmost panels display the posterior distribution over all the possible combinations (*k, m*), with the red marker indicating the maximum a posteriori value.

**Extended Data Figure 2.**
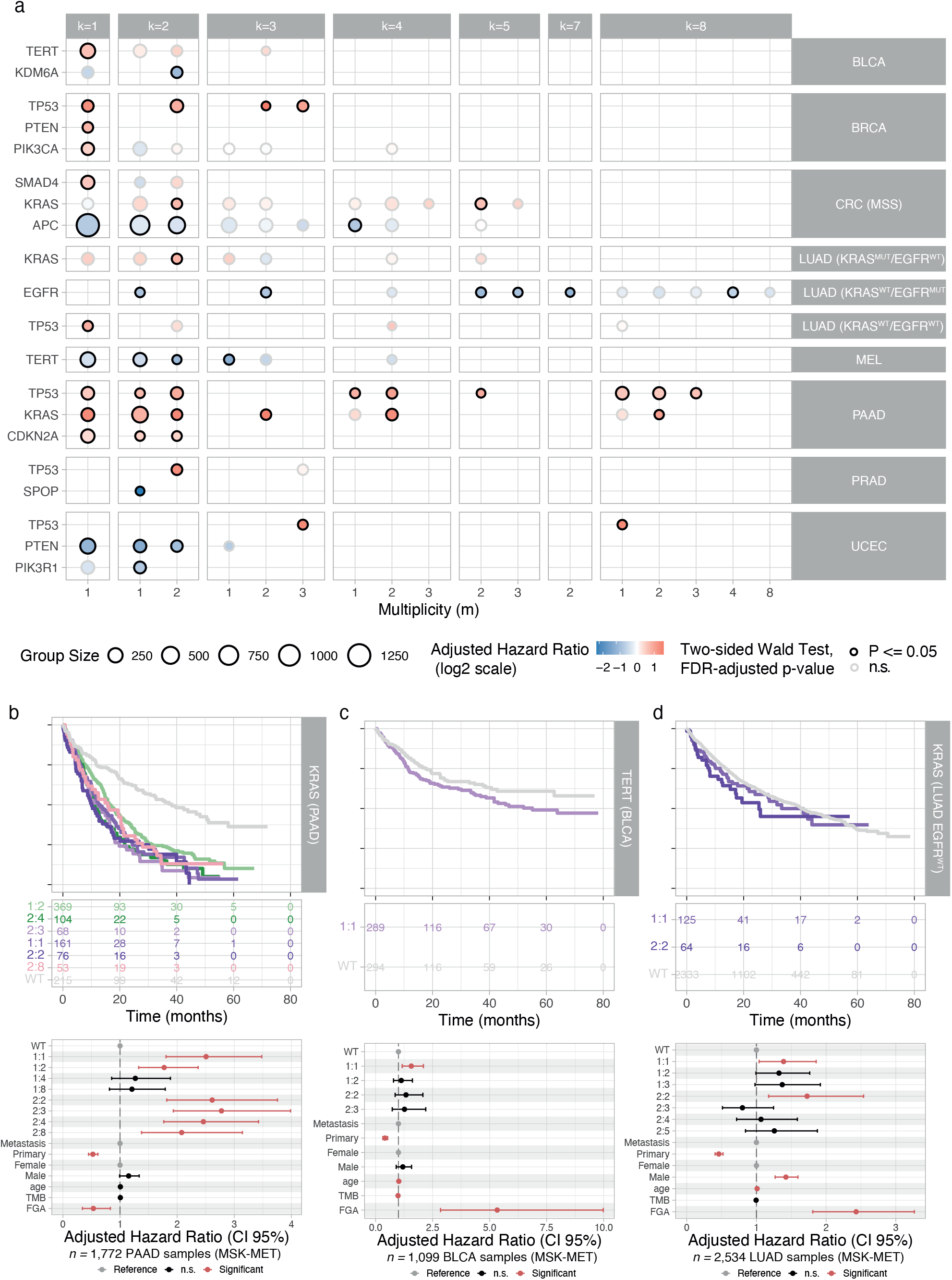
Pan-cancer survival analyses at mutation multiplicity and copy number resolution. a. Cox multivariate regression, using sample type (primary vs metastasis), sex, age, TMB and FGA as covariates, at (*k, m*) resolution for MSK-MET patients. Survival analysis, including Kaplan-Meier estimation and Cox regression hazard ratio estimates for b. KRAS mutant pancreatic adenocarcinoma, c. TERT mutant bladder cancer, d. KRAS mutant, EGFR-WT lung adenocarcinoma. Groups are named by (*k* : *m*), with wild-type (WT) patients as reference. n.s: non-significant.

**Extended Data Figure 3.**
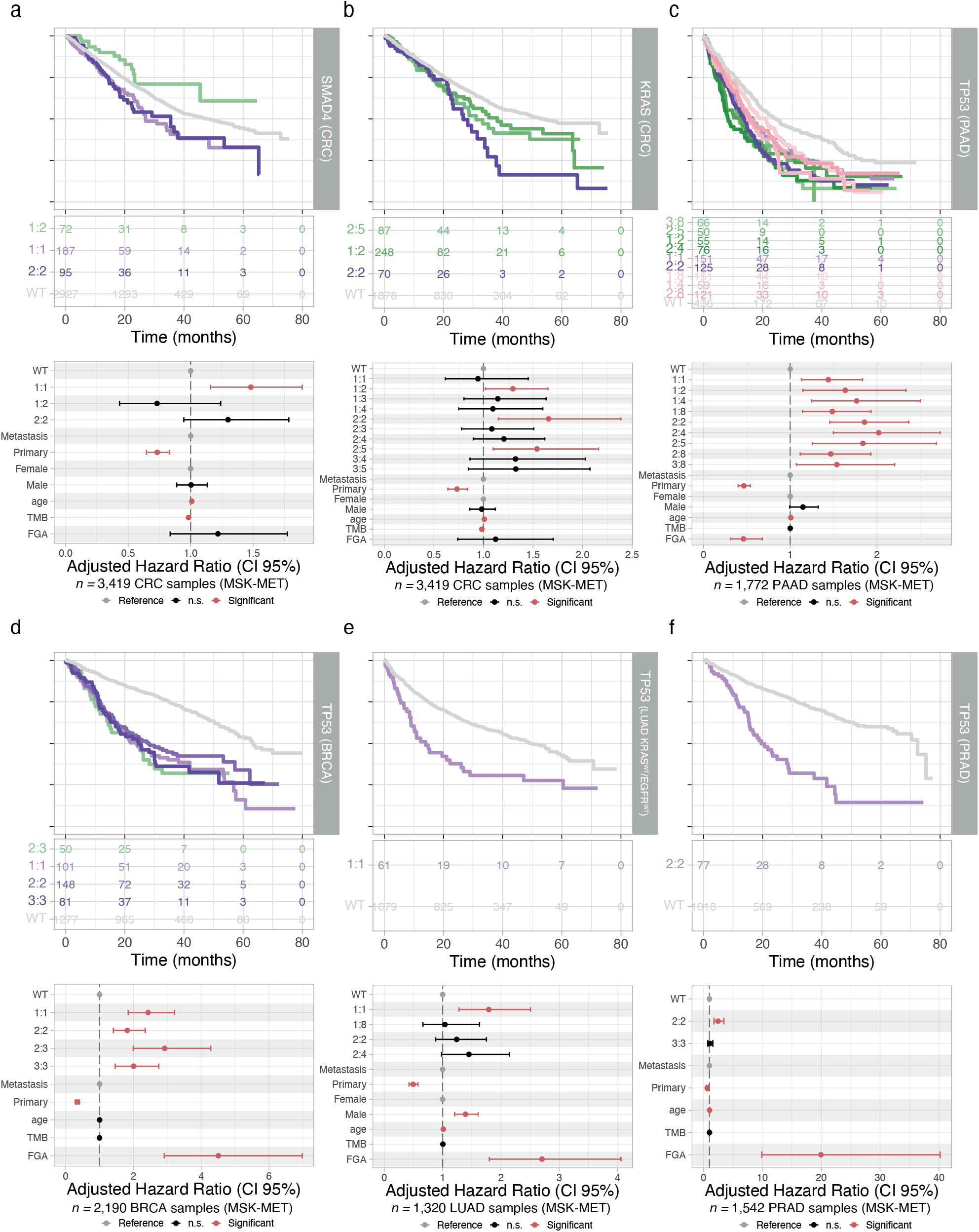
Survival analysis of MSK-MET samples at full allelic resolution. Overall survival analysis including Kaplan-Meier estimation and Cox multivariate regression, using sample type (primary vs metastasis), sex, age, TMB and FGA as covariates, at (*k, m*) resolution for MSK-MET patients with a. SMAD4 mutant colorectal cancer, b. KRAS mutant colorectal cancer, c. TP53 mutant pancreatic adenocarcinoma, d. TP53 mutant breast cancer, e. TP53 mutant lung cancer with no KRAS or EGFR mutations, and f. TP53 mutant prostate cancer. Groups are named by (*k* : *m*), with wild-type (WT) patients as reference. n.s: non-significant.

**Extended Data Figure 4.**
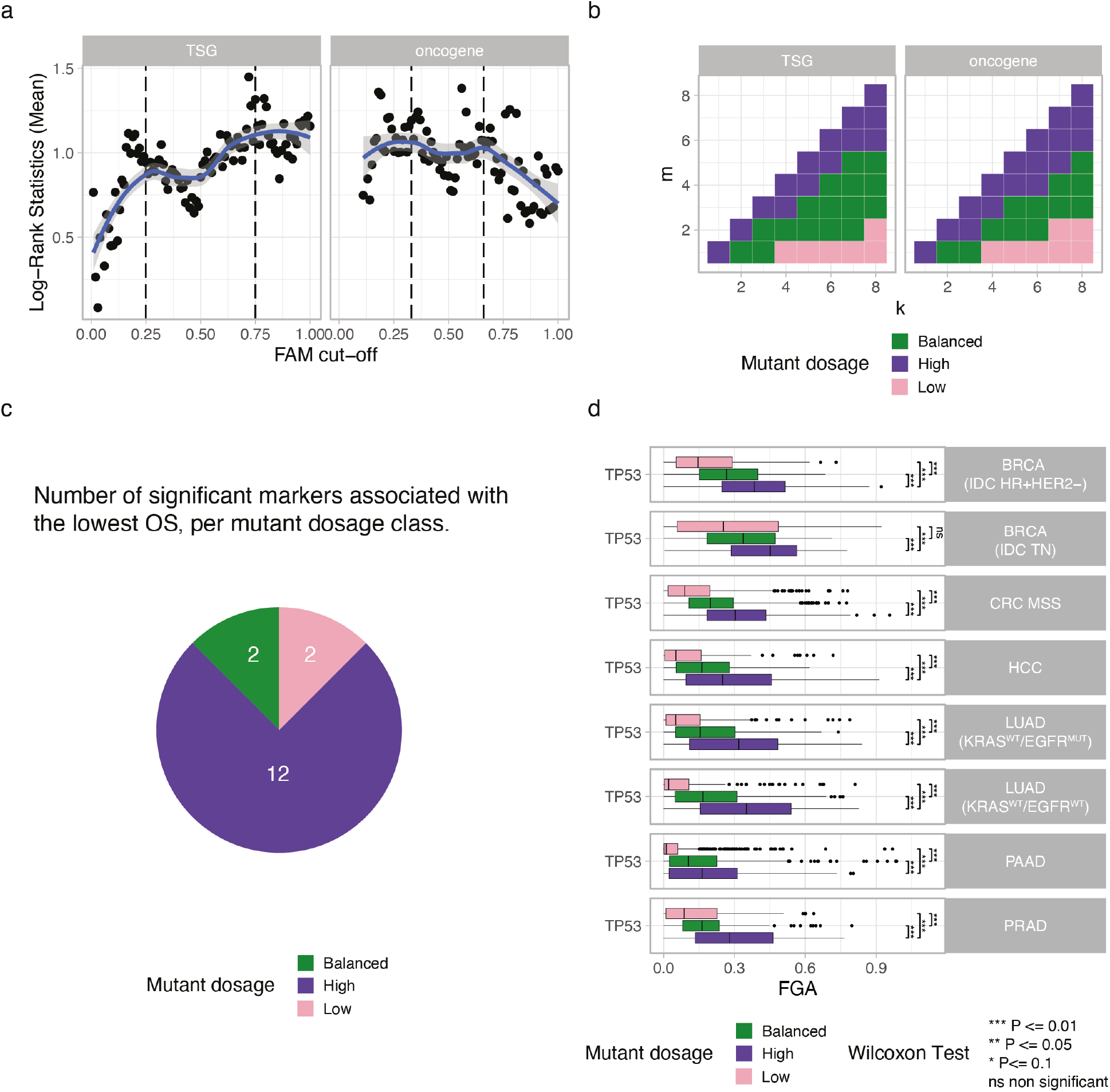
Definition and characterization of gene mutant dosage classes. a. Maximally selected rank statistics quantifies the optimal lower and upper cut-points of the fraction of allele with the mutation (FAM) in overall survival (OS) analysis stratifying mutant vs wild-type patients across 84 tumor suppressor genes (TSG, optimal values 25% and 75%), 35 oncogenes (optimal values 33% and 66%) and 18 tumour types. b. Classification of mutation total copy number and multiplicity combinations (*k, m*) into Low, Balanced and High mutant dosage classes. c. Relative fraction of prognostic gene mutant dosage classes across significant markers. d. Distribution of the fraction of genome altered (FGA) across gene mutant dosage classes of TP53 in tumour types, compared through pairwise Wilcoxon test.

**Extended Data Figure 5.**
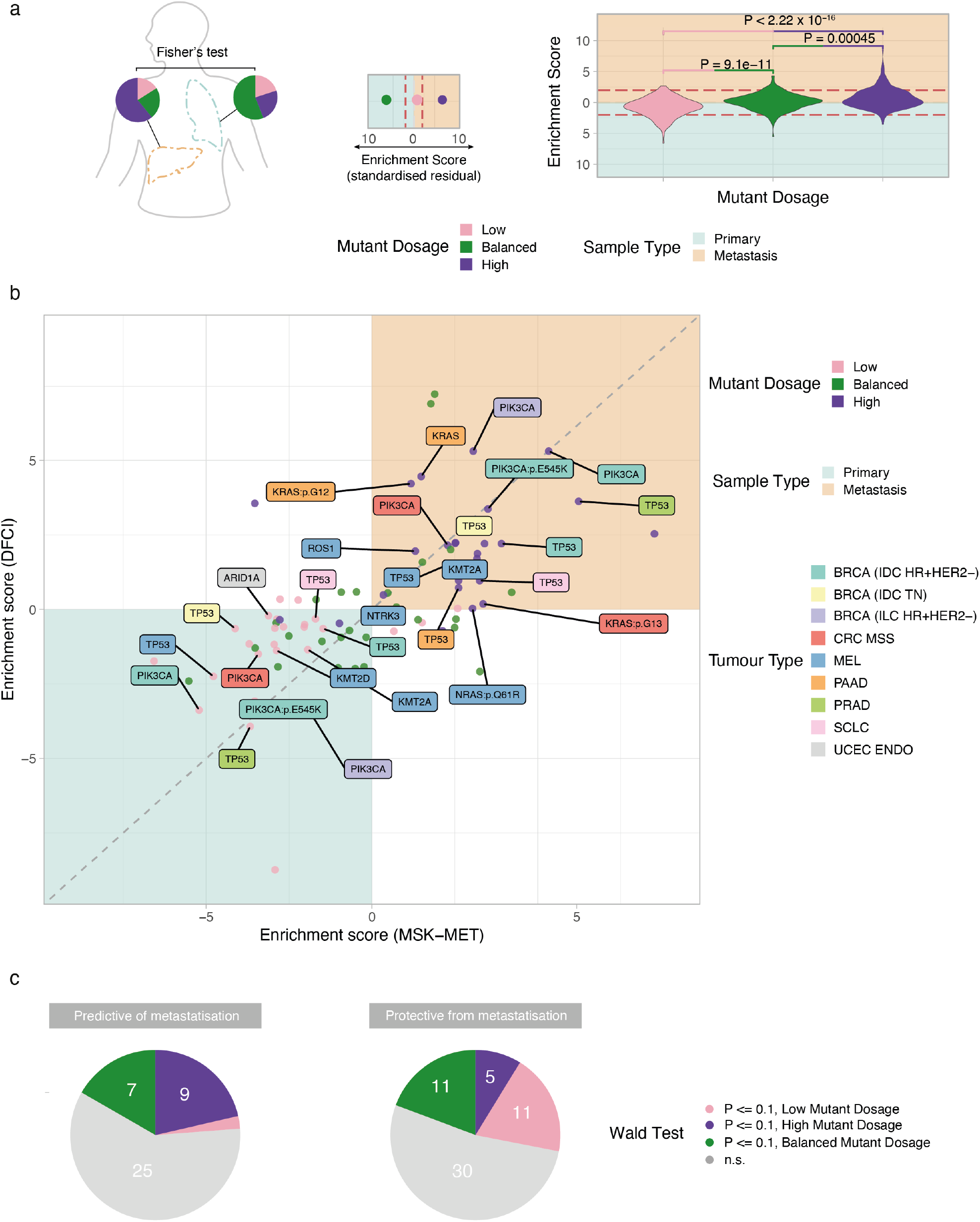
Consistency and distribution of gene mutant dosage enrichment and metastatic associations. a. Distribution of enrichment scores in primary tumours vs metastases, defined as the standardised residuals of Fisher’s exact test, across mutant dosage classes. b. Comparison of enrichment scores between the MSK-MET and the DFCI cohorts. Significant markers with consistent enrichment scores (distance lower than independent expected noise) across the two cohorts are highlighted. c. Distribution of mutant gene dosage classes across markers of increased (left) and decreased (right) metastatic propensity using a univariate logistic model.

## 7. Supplementary Information

### 7.1. Interpretation of posterior predictive checks and sparse-mutation regimes

After estimating the full posterior distribution of model parameters Θ = (*π, η, k, m*), INCOMMON evaluates the consistency between observed data and model-implied distributions for total copy number *k*, multiplicity *m*, per-copy read rate *η*, and tumour purity *π* using posterior predictive checks (PPCs). Rather than acting as binary acceptance/rejection criteria, PPCs provide structured information about where and how discrepancies arise. Within the MSK-MET cohort, in addition to perfectly fitted samples (*N* = 14259), we identified the following three major scenarios:

#### 7.1.1. Localised discrepancies

In *N* = 6631 samples, although most parameters were well calibrated, we observed minor deviations, including a posterior purity estimate that deviated slightly from the prior mean. In such cases, the discrepancy typically propagated to localised copy-number errors, such as single or a few mutations whose observed variant reads fell in the tail of the posterior predictive distribution (e.g. Supplementary Figure S5).

These situations may reflect locus-specific technical variation, mild subclonality, or prior misspecification, but they do not necessarily indicate model failure at the sample level. INCOMMON explicitly reports these deviations, allowing users to distinguish between isolated events and systemic misfit.

#### 7.1.2. Global instability

By contrast, in a smaller number of samples (*N* = 1076), PPC failures were coordinated across multiple model parameters (e.g. Supplementary Figure S6). Such discrepancies suggested a structural incompatibility between observed data and the model assumptions, potentially due to inaccurate purity input, unmodelled tumour heterogeneity, or overly complex copy-number architectures. In these cases, INCOMMON flags the sample as poorly fitted, preventing unreliable estimates from entering downstream analyses.

#### 7.1.3. Inference with one or two mutations: regularisation and uncertainty

Targeted panel assays frequently yield samples with only one or two detectable mutations. When a single mutation is internally consistent with the joint constraints of the model (e.g. Supplementary Figure S7), INCOMMON can still infer tumour purity, per-copy read rate, total copy number, and multiplicity. The Bayesian formulation was critical in this context: priors on *π* and *η* provided regularisation, preventing degenerate solutions and yielding biologically plausible posterior distributions. In such cases, posterior predictive p-values for the Poisson depth model may approach 1 because the inferred copy number closely matches the observed depth. This behaviour reflected limited information rather than overfitting or model misspecification. Importantly, uncertainty was appropriately reflected in broader posterior distributions.

Conversely, when even a single mutation was inconsistent with the joint constraints imposed by sequencing depth and sample purity (e.g. Supplementary Figure S8), PPCs revealed simultaneous deviations in total copy number, multiplicity, and purity. This demonstrated that sparsity alone does not explain poor fit; rather, inconsistency between the observed data and the model’s generative structure drives the fit diagnostics.

Thus, INCOMMON can both stabilise inference in underdetermined settings and detect incompatibility in minimal-data scenarios.

### 7.2. Comparative evaluation of INCOMMON mutant dosage and purity-adjusted VAF stratification

To further clarify the added value of INCOMMON relative to tumour purity-adjusted VAF (VAF/*ρ*), we performed a systematic comparison of the two approaches across univariate and multivariate Cox regression frameworks, assessed overlap in significant discoveries, evaluated robustness to purity misspecification, and examined biological plausibility of uniquely identified associations.

#### 7.2.1. Overlap and differential discoveries

Across tumour types, the adjusted VAF approach identified 248 significant associations in univariate models and 121 in multivariate models (FDR < 0.1), compared to 260 and 147, respectively, for INCOMMON mutant dosage. Thus, INCOMMON yielded +12 additional significant classes in univariate analysis and +26 in multivariate analysis.

A comprehensive comparison, including hazard ratios (mean and 95% CI) and FDR-adjusted p-values for both approaches, is provided in Supplementary Table S10. The majority of associations identified by adjusted VAF were also detected by INCOMMON, whereas a non-trivial subset of robust multivariate associations were uniquely recovered by mutant dosage.

#### 7.2.2. Biological support for associations uniquely detected by INCOMMON

Importantly, several associations uniquely identified by INCOMMON in multivariate analysis are strongly supported by independent biological and clinical evidence. In breast cancer, mutant dosage uniquely recovered associations involving *CDH1* (Balanced; HR = 1.56, CI = 1.18–2.06, *P* = 0.0133), *GATA3* (Balanced; HR = 0.708, CI = 0.553– 0.906, *P* = 0.0423), and *PIK3CA* (Low; HR = 0.628, CI = 0.476–0.827, *P* = 0.0133), all of which have well-established prognostic or subtype-defining roles^52,53,56^.

In colorectal cancer, INCOMMON uniquely detected robust effects for *APC* (High class; HR = 0.647, CI = 0.561– 0.746, *P* < 0.0001), *BRAF* (High class; HR = 2.17, CI = 1.46–3.23, *P* = 0.00609; Low class; HR = 1.84, CI = 1.30– 2.60, *P* = 0.0180), *CTCF* (High class; HR = 10.6, CI = 2.64–42.9, *P* = 0.0230), *FAT1* (Intermediate class; HR = 0.469, CI = 0.303–0.727, *P* = 0.0203), *MAP3K1* (High class; HR = 2.82, CI = 1.46–5.45, *P* = 0.0450), *SMAD4* (High class; HR = 1.48, CI = 1.22–1.79, *P* = 0.00423), and *TCF7L2* (Low class; HR = 0.521, CI = 0.361–0.751, *P* = 0.0180), each previously implicated in tumour progression, chromosomal instability, WNT signalling, or clinical outcome^24,45–47,55,58,98.^

In hepatocellular carcinoma, dosage-based stratification identified prognostic effects for *TP53* (Low class; HR = 1.52, CI = 1.14–2.02, *P* = 0.0142), consistent with established *TP53*-associated immune and survival models^59^.

In lung adenocarcinoma, INCOMMON uniquely detected associations for *EGFR* (High class; HR = 0.700, CI = 0.577– 0.848, *P* = 0.00411; Low class; HR = 0.647, CI = 0.553–0.756, *P* < 0.0001), *KEAP1* (Low class; HR = 1.76, CI = 1.35–2.29, *P* = 0.000581), *SMARCA4* (High class; HR = 2.00, CI = 1.51–2.66, *P* < 0.0001; Low class; HR = 1.71, CI = 1.29–2.26, *P* = 0.00295), and *TBX3* (Low class; HR = 0.280, CI = 0.125–0.627, *P* = 0.0230), all supported by prior evidence linking dosage or mutation context to treatment response and survival^44,48,49,60^.

In pancreatic adenocarcinoma, mutant dosage stratification highlighted effects for *ARID1A* (Intermediate class; HR = 1.35, CI = 1.03–1.77, *P* = 0.0496), *CDKN2A* (High class; HR = 1.26, CI = 1.11–1.43, *P* = 0.000695), *RNF43* (High class; HR = 0.521, CI = 0.293–0.925, *P* = 0.0496), and *TP53* (Low class; HR = 1.59, CI = 1.32–1.90, *P* < 0.0001), consistent with previous clinicogenomic studies implicating these drivers in pancreatic tumour progression and outcome^14–16^.

Finally, in uterine cancer, additional robust associations were identified for *ARID1A* (Intermediate class; HR = 0.657, CI = 0.490–0.879, *P* = 0.0489), *BCOR* (Low class; HR = 0.116, CI = 0.0285–0.471, *P* = 0.0343), *ERBB3* (High class; HR = 4.14, CI = 1.53–11.2, *P* = 0.0490), and *PTEN* (High class; HR = 0.507, CI = 0.381–0.674, *P* < 0.0001), all consistent with known endometrial tumorigenesis pathways^50,51,54,99^.

Collectively, these results indicate that the additional associations detected by INCOMMON are not random or unstable findings, but instead correspond to biologically coherent and independently supported potential prognostic relationships.

#### 7.2.3. Precision of hazard ratio estimates

Beyond differences in the number of significant discoveries, Cox regression analyses demonstrated systematically wider 95% confidence intervals for hazard ratios derived from adjusted VAF stratification (Supplementary Figure S10d), with INCOMMON-based mutant dosage stratification yielding, on average, a 16.93% smaller standard deviation of hazard ratio estimates, on average. Consistently, in both univariate and multivariate models, the variability of hazard ratio estimates was generally higher for adjusted VAF than for INCOMMON (Supplementary Figure S10e). The largest differences were observed in the high-dosage class (FDR-adjusted p-value under t-test *P* = 3.68 × 10^−16^ for univariate models; *P* = 6.12 × 10^−17^ for multivariate models), where adjusted VAF exhibited substantially higher median standard errors (0.504 and 0.579 for univariate and multivariate models, respectively) compared with mutant dosage (0.318 and 0.321), corresponding to nearly twofold greater variability. Differences were considerably smaller in the other dosage classes. In the low-dosage class, adjusted VAF showed modestly higher median standard errors (0.335 and 0.343 for univariate and multivariate models, respectively; FDR-adjusted *P* = 2.81 × 10^−2^ and *P* = 1.79 × 10^−2^) than mutant dosage (0.295 and 0.301). In the intermediate-dosage class, the two approaches produced very similar estimates, with adjusted VAF showing slightly lower median standard errors (0.174 and 0.182; FDR-adjusted *P* = 3.05 × 10^−5^ and *P* = 3.46 × 10^−5^) than mutant dosage (0.202 and 0.210). Overall, these results indicate that even when both approaches detect significant associations, mutant dosage generally provides more precise and stable estimates of effect size, with the most pronounced improvements observed for high-dosage mutations.

#### 7.2.4. Analytical distinction between adjusted VAF and FAM

Although purity-adjusted VAF may approximate the fraction of alleles carrying the mutation (FAM) in specific configurations, the two quantities are analytically distinct. The expected VAF conditional on tumour purity, mutation multiplicity (*m*), and total copy number (*k*) depends jointly on all three parameters. The adjusted VAF equals FAM only under restricted purity and copy-number regimes. In particular, taking limits over tumour purity shows that (i) at high purity, adjusted VAF systematically overestimates FAM (converging to *m*/*k* but remaining biased in finite samples), and (ii) at low purity, adjusted VAF converges to approximately *m*/2 only when *k* = 1 and diverges for other copy-number states.

Thus, adjusted VAF does not provide a consistent estimator of the fraction of mutant alleles across copy-number contexts. In contrast, INCOMMON explicitly models mutation multiplicity and total copy number, directly targeting the biologically interpretable quantity of mutant allele dosage.

#### 7.2.5. Sensitivity to tumour purity misspecification

We next examined the impact of tumour purity misspecification through simulation. We generated 1,000 tumour samples across varying purity levels and mutation burdens, sampling copy number and mutation multiplicity from prior distributions derived from PCAWG and HMF. Total and variant reads were simulated from Poisson and Binomial distributions, respectively. Simulated purities were then perturbed with errors ranging from 0–100% (median 12.7%, matching TCGA consensus pipeline estimates), enabling direct comparison against ground-truth FAM. Under increasing purity error, the adjusted VAF approach exhibited marked degradation in accuracy, with FAM error rising to ∼ 50%. In contrast, INCOMMON remained comparatively stable, with error remaining below 20% across the same perturbation range. Quantitatively, adjusted VAF accumulated approximately 10% additional FAM error for every 10% absolute increase in purity error relative to INCOMMON.

This behaviour is consistent with the deterministic structure of adjusted VAF, which divides observed VAF by a fixed purity estimate and therefore directly propagates misspecification error. In contrast, INCOMMON adopts a Bayesian framework that jointly models purity, copy number, and mutation multiplicity, integrating uncertainty into posterior inference and attenuating downstream bias.

### 7.3. Supplementary Tables

- Supplementary Table S1: Priors on (*k, m*) from WGS data
- Supplementary Table S2: Priors on count rate per copy
- Supplementary Table S3: Computational efficiency
- Supplementary Table S4: Survival analysis (KM)
- Supplementary Table S5: Survival analysis (Cox)
- Supplementary Table S6: Mutant Dosage vs Adjusted VAF
- Supplementary Table S7: Survival analysis at (*k, m*) resolution
- Supplementary Table S8: Mutant Dosage enrichment
- Supplementary Table S9: Metastatic propensity
- Supplementary Table S10: Metastatic tropism
- Supplementary Table S11: HMF to INTOGEN conversion

### 7.4. Supplementary Figures

**Supplementary Figure S1.**
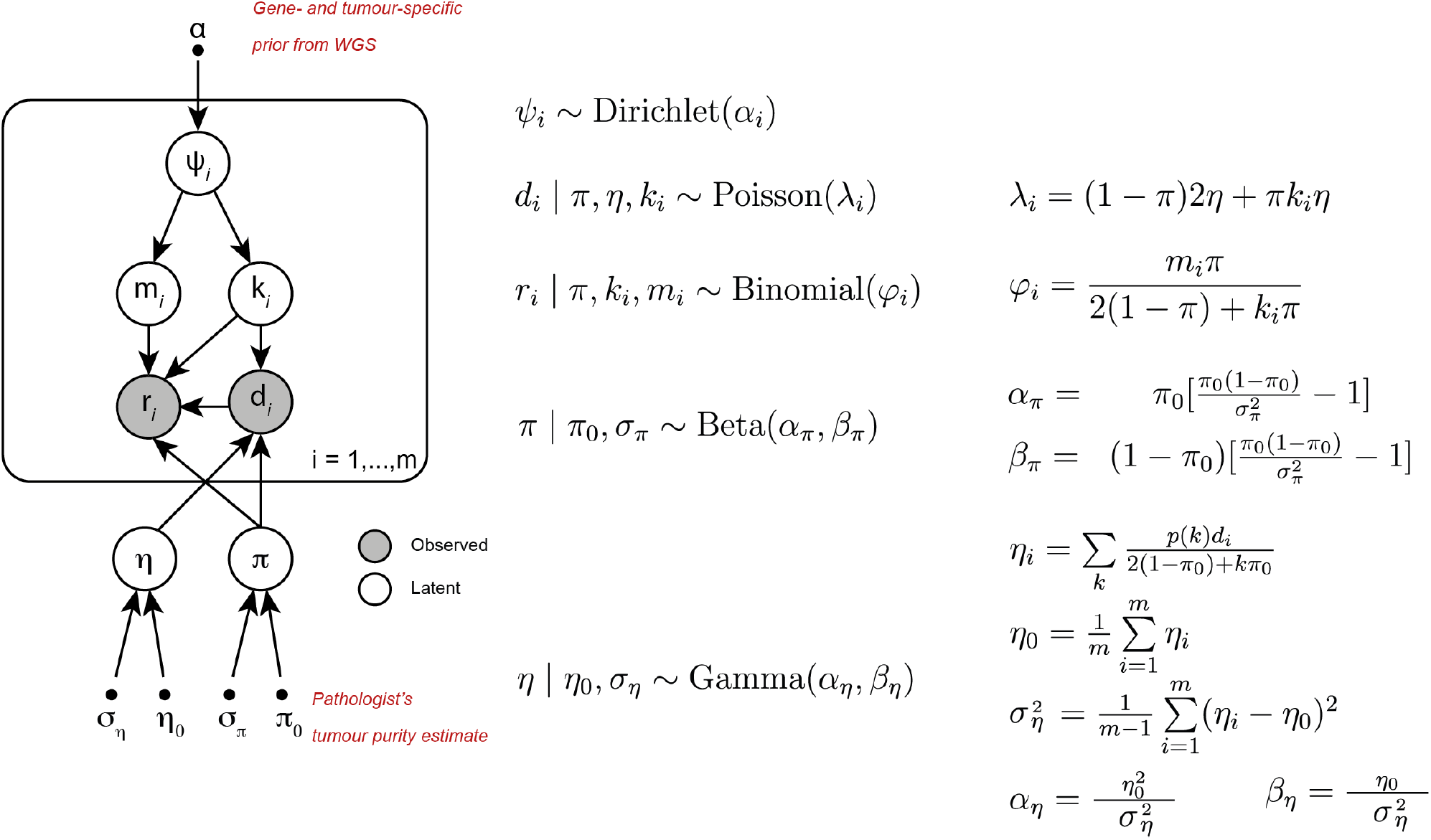
Probabilistic graphical model of INCOMMON. Unobserved (latent) variables are indicated as white circles, observed variables as grey circles and fixed hyperparameters as dots. Prior probabilities *ψ* for each combination of mutation total copy number *k* and multiplicity *m* are sampled from a Dirichlet distribution, with concentration parameters *α* reflecting the relative fraction observed from large-scale WGS cohorts. The prior probability for tumour purity *π* is centred at the pathologist’s estimate *π*_0_ with fixed variance *σ*_*π*_, whereas the mean *η*_0_ and variance *σ*_*η*_ of the prior probability for the read count rate per chromosome copy *η* are estimated from the data. The total number of reads at each site *d* follows a Poisson distribution with rate *λ* contributed by normal ((1 − *π*)2*η*) and tumour (*πkη*) cells. The number of reads with the variant *r* follows a Binomial distribution with success probability *ϕ* defined as the expected variant allele frequency *mπ*/(2(1 − *π*) + *kπ*).

**Supplementary Figure S2.**
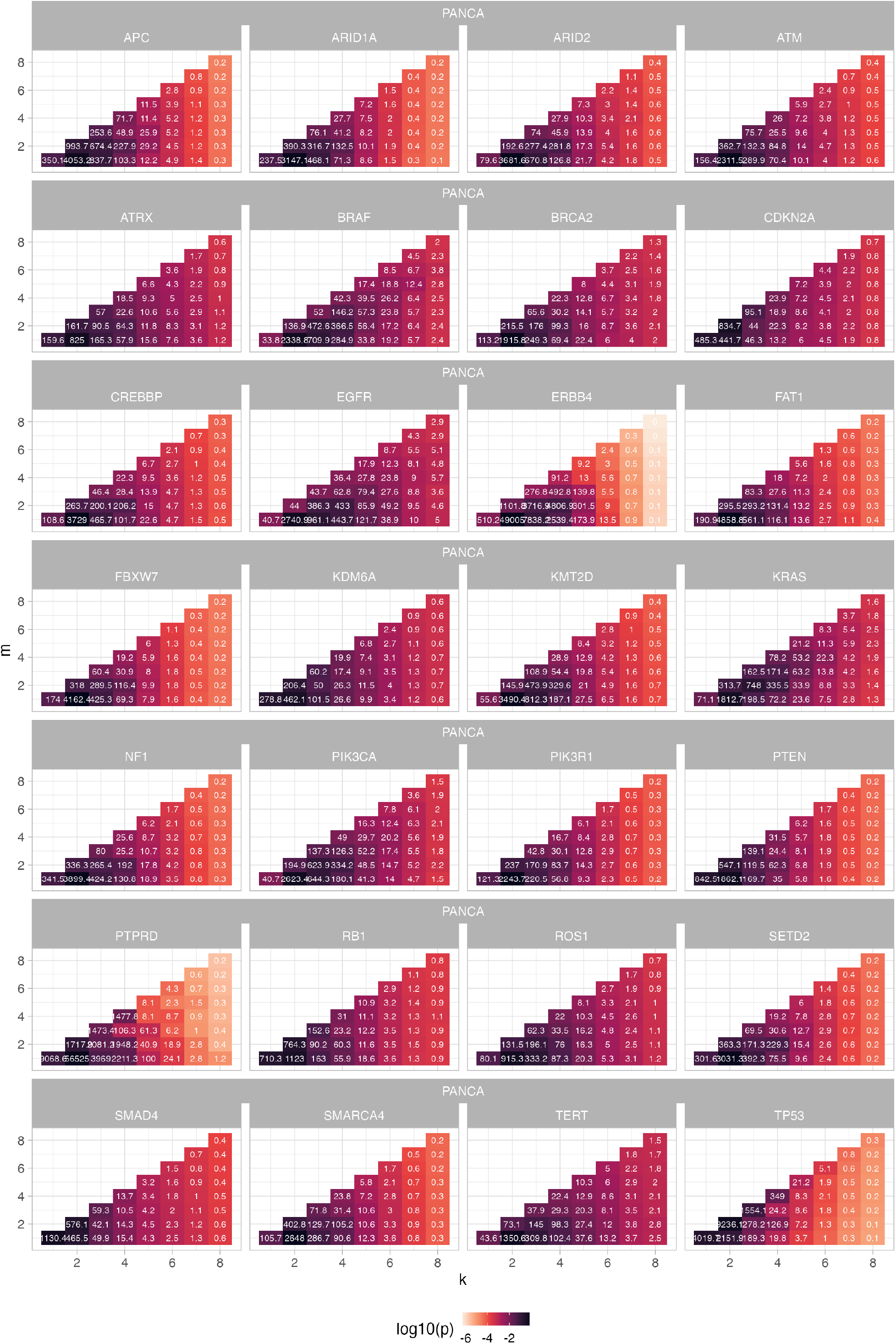
Pan-cancer (PANCA) prior distribution on the copy number and multiplicity configurations (*k, m*) obtained from the PCAWG and HMF whole genome sequencing cohorts. The color of each cell corresponds to the relative fraction of the configuration in the dataset, while the number label inside is equal to the actual count (or pseudo-count, see Methods), used as Dirichlet concentration parameter.

**Supplementary Figure S3.**
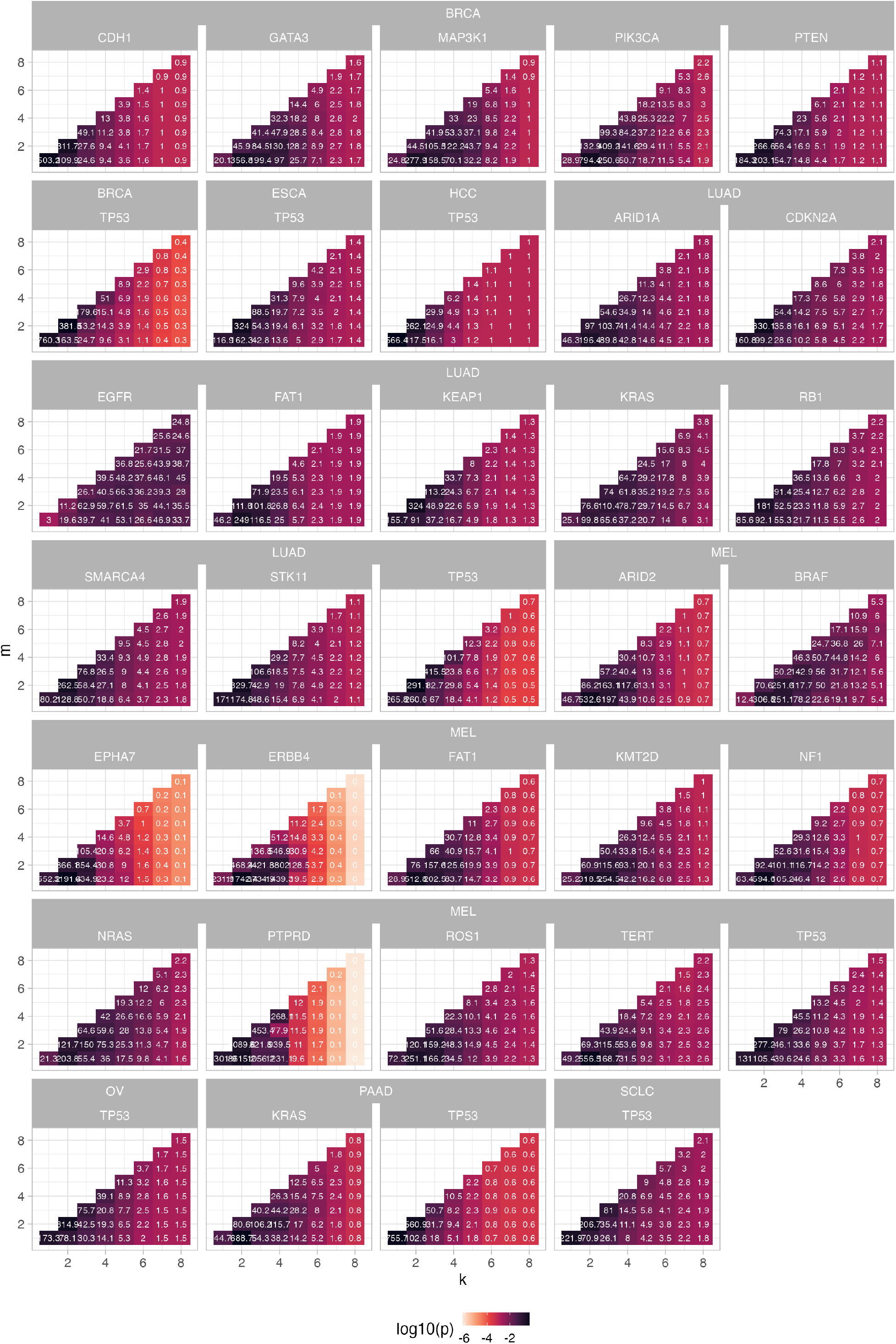
Tumour type-specific prior distribution on the copy number and multiplicity configurations (*k, m*) obtained from the PCAWG and HMF whole genome sequencing cohorts. The color of each cell corresponds to the relative fraction of the configuration in the dataset, while the number label inside is equal to the actual count (or pseudo-count, see Methods), used as Dirichlet concentration parameter.

**Supplementary Figure S4.**
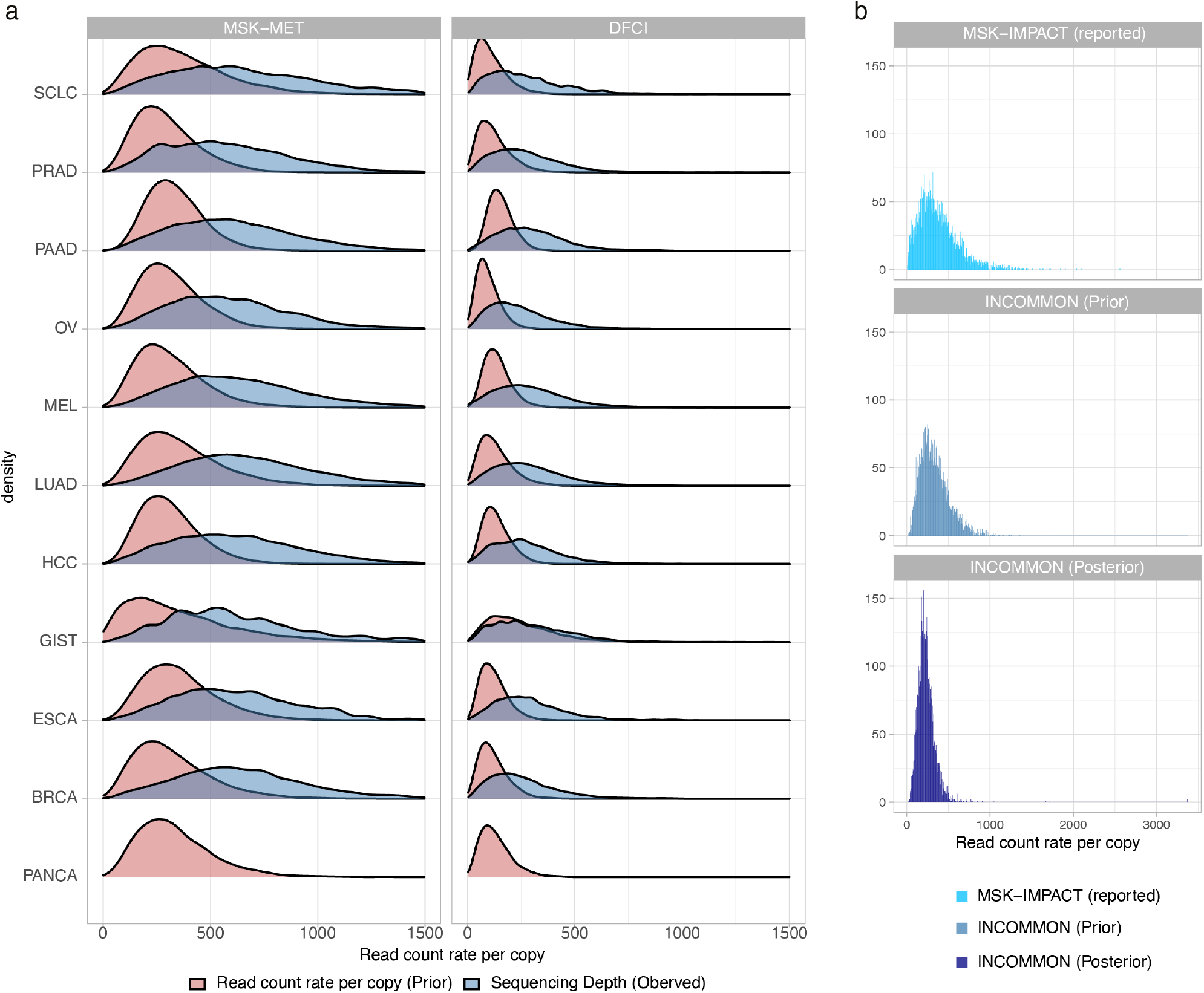
a. Gamma prior distribution (red) over the read count per chromosome copy, compared with the observed distribution of sequencing depth (blue), across the MSK-MET and GENIE-DFCI cohorts. Notice that the empirical distribution of sequencing depth is confounded by mutation copy number and tumour purity. b. Read count rate per chromosome copy as reported for the MSK-IMPACT targeted panel[21], aligning with prior and posterior INCOMMON distributions.

**Supplementary Figure S5.**
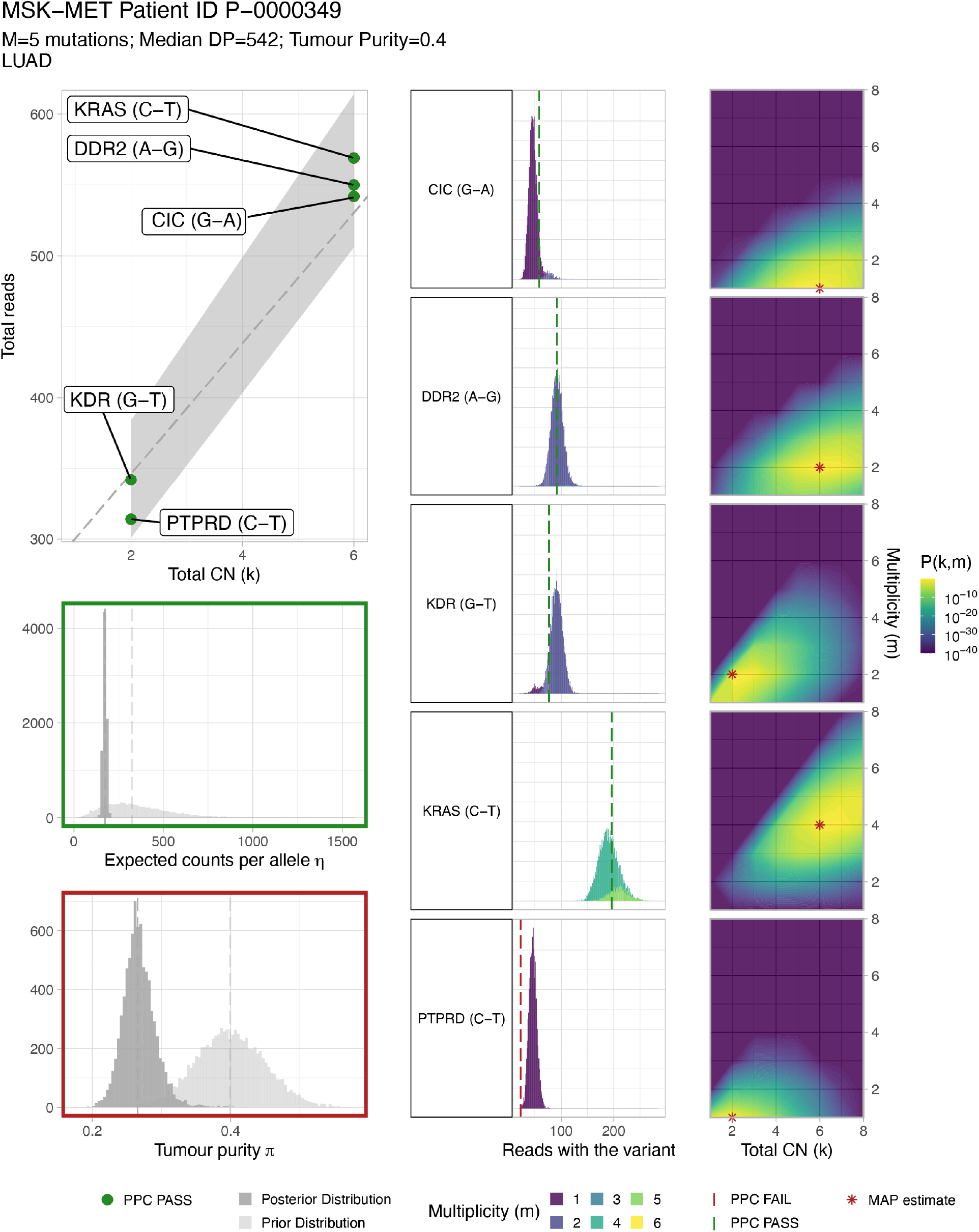
INCOMMON report for MSK-MET lung cancer patient P-0000349. All posterior predictive checks (PPC) are passed, except for tumour purity (99.4% of samples from the posterior distribution - mean *π* = 0.251 - deviate from the prior mean *π*_0_ = 0.4 more than expected a priori) and multiplicity of the C-T mutation on PTPRD gene (the observed number of reads with the variant *NV* = 23 lies in the lower 5% tail of the posterior predictive distribution, indicating a poor fit).

**Supplementary Figure S6.**
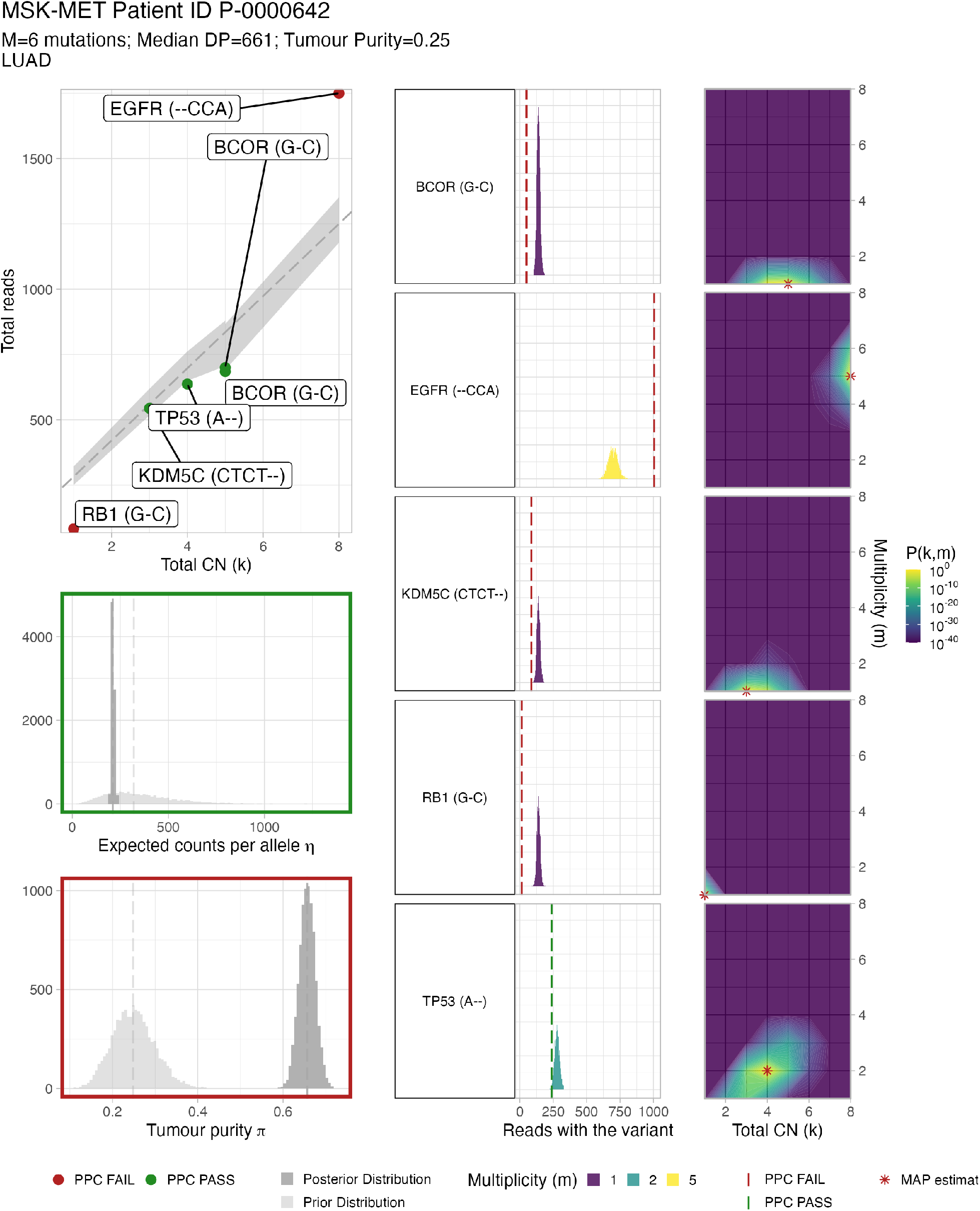
INCOMMON report for MSK-MET lung cancer patient P-0000642, a case of poor fit, failing posterior-predictive checks (PPC) on total copy number for G-C substitution on RB1 and CCA insertion on EGFR, tumour purity (inferred *π* = 0.641 versus prior estimate *π*_0_ = 0.25, with no overlap between prior and posterior distributions), multiplicity of on all sequenced alterations except for an A deletion on TP53.

**Supplementary Figure S7.**
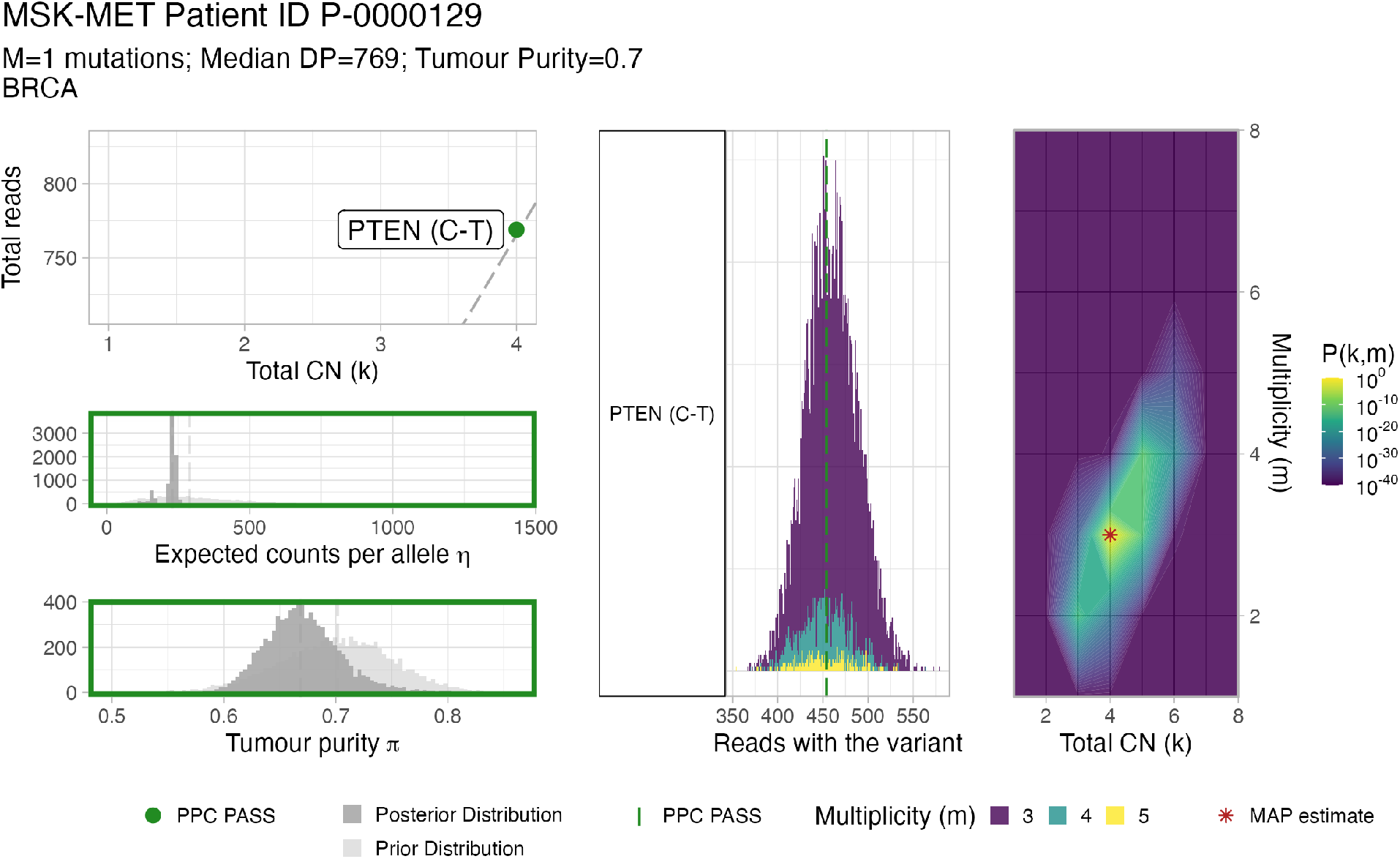
INCOMMON report for MSK-MET breast cancer patient P-0000129, a particular case with one single mutation, a C-T substitution in the PTEN gene, is detected by the targeted panel assay. Even if one single data point is available, INCOMMON is able to infer the expected read counts per allele *η*, the tumour purity *π*, and the mutation total copy number and multiplicity (*k* = 4, *m* = 3), in this case. The inference is feasible thanks to the Bayesian formulation of INCOMMON, which allows leveraging prior distributions for the model parameters Θ = (*π, η, k, m*).

**Supplementary Figure S8.**
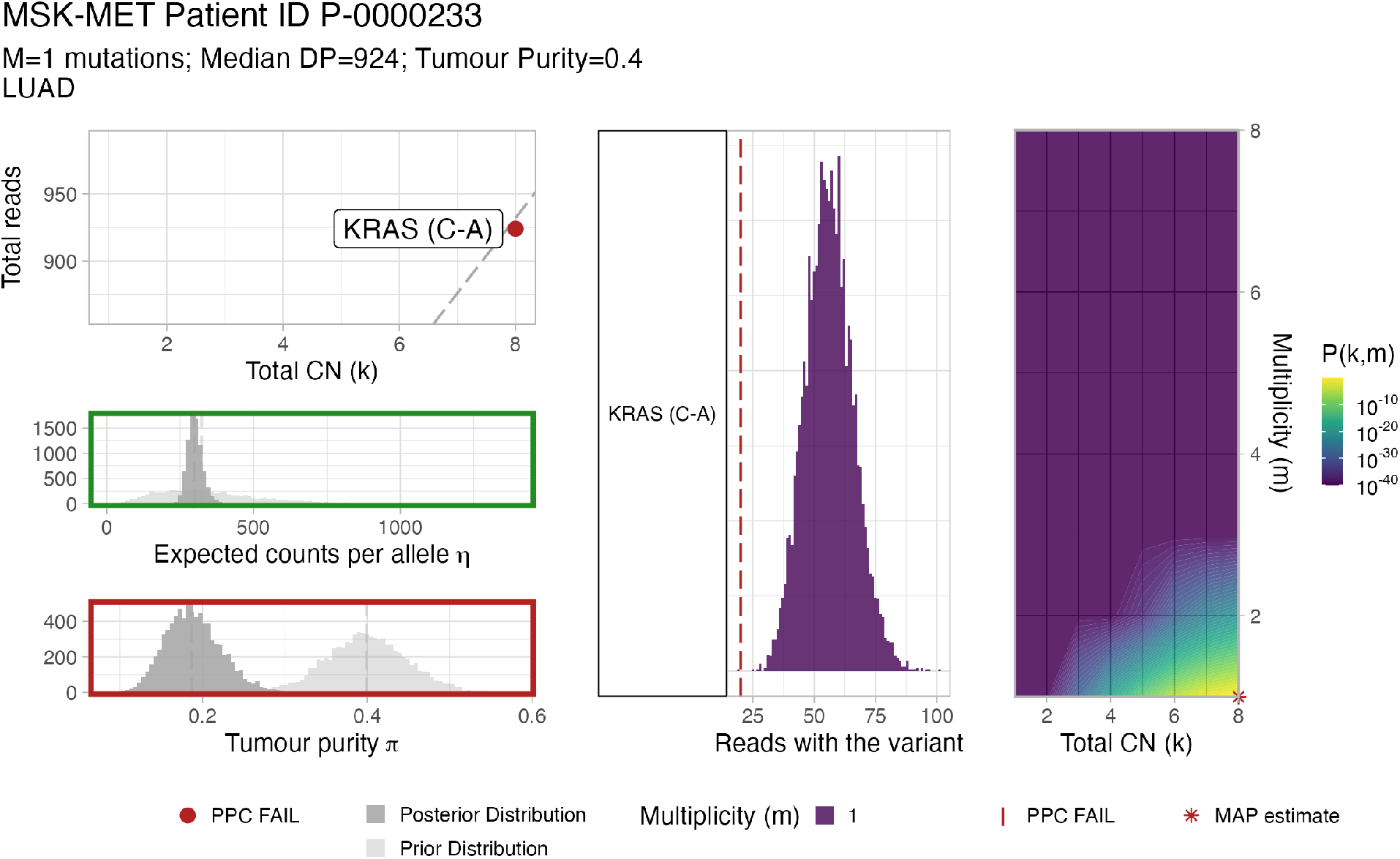
INCOMMON report for MSK-MET lung cancer patient P-0000233, a case with one single mutation, a C-A substitution in the KRAS gene, is detected by the targeted panel assay. In this particular case, posterior-predictive checks indicate a poor fit of total copy-number (total number of reads lower than expected), tumour purity (deviation from prior input purity outside the allowed error of *ε* = ±10%15%), and multiplicity (number of reads with the variant lower than expected). This demonstrates the capability of INCOMMON to flag samples with a poor fit.

**Supplementary Figure S9.**
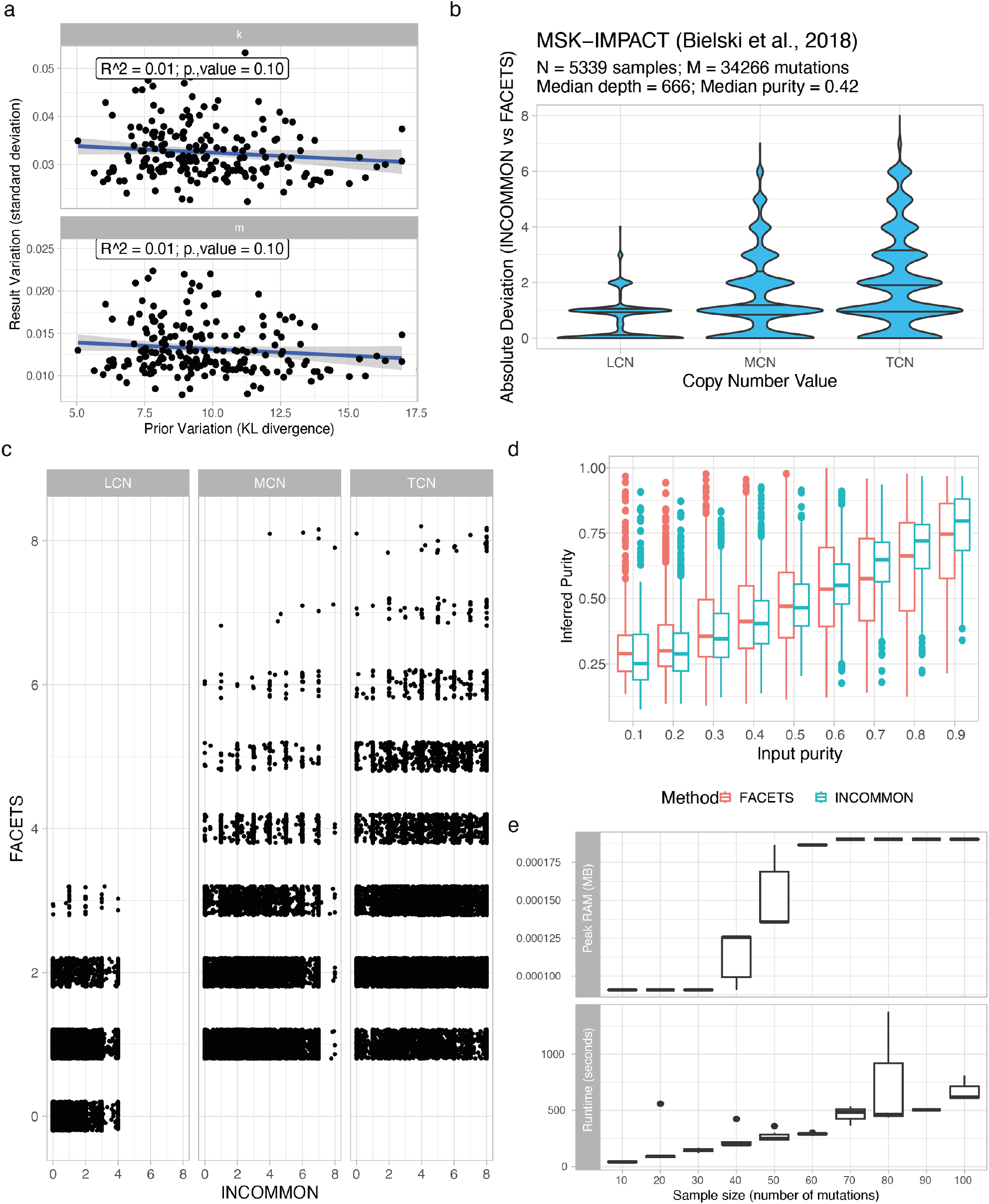
a. Standard deviation of the mean absolute error (MAE) estimates against prior variation measured by Kullback-Leibler divergence, across *N* = 50 iterations of random-splitting (70% training, 30% test) of the combined PCAWG and HMF data. b. Comparison of INCOMMON and FACETS copy-number calls across the MSK-IMPACT dataset published in Bielski et. al.^33^ c. Comparison of INCOMMON and FACETS inference of tumour purity. d. The low peak RAM usage (top panel) and runtime required for the classification of samples of increasing size (number of mutations) demonstrate the high computational efficiency of INCOMMON.

**Supplementary Figure S10.**
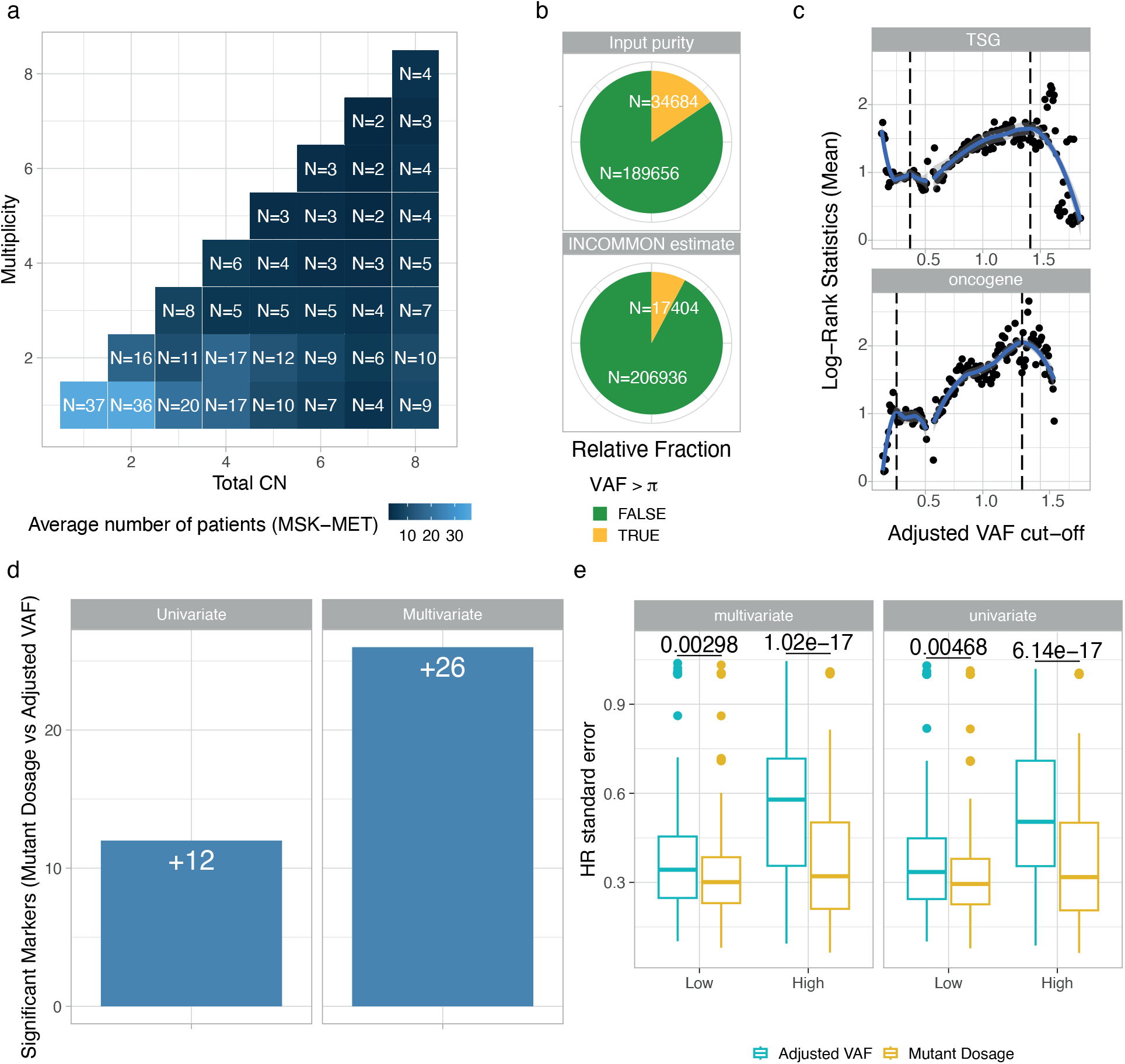
a. Average number of patients per (*k, m*) configuration in the MSK-MET cohort, across genes and tumour types. b. The relative fraction of MSK-MET samples with inconsistent VAF and tumour purity (VAF > *π*) decreases when using the INCOMMON estimate instead of the publicly available input purity. c. Optimal adjusted VAF (VAF/*π*) thresholds for patient stratification into Low, Intermediate and High adjusted VAF, maximising the average Log-Rank statistics across TSGs, oncogenes and tumour types in the MSK-MET cohort. d. Absolute increase in the number of significant Mutant Dosage markers with respect to adjusted VAF classification, in univariate and multivariate Cox regression (using patients’ age, sex, TMB, FGA and sample type as covariates). e. The standard error of hazard ratios (HR) in multivariate Cox regression, using adjusted VAF stratification, is significantly higher than in Mutant Dosage stratification.

**Supplementary Figure S11.**
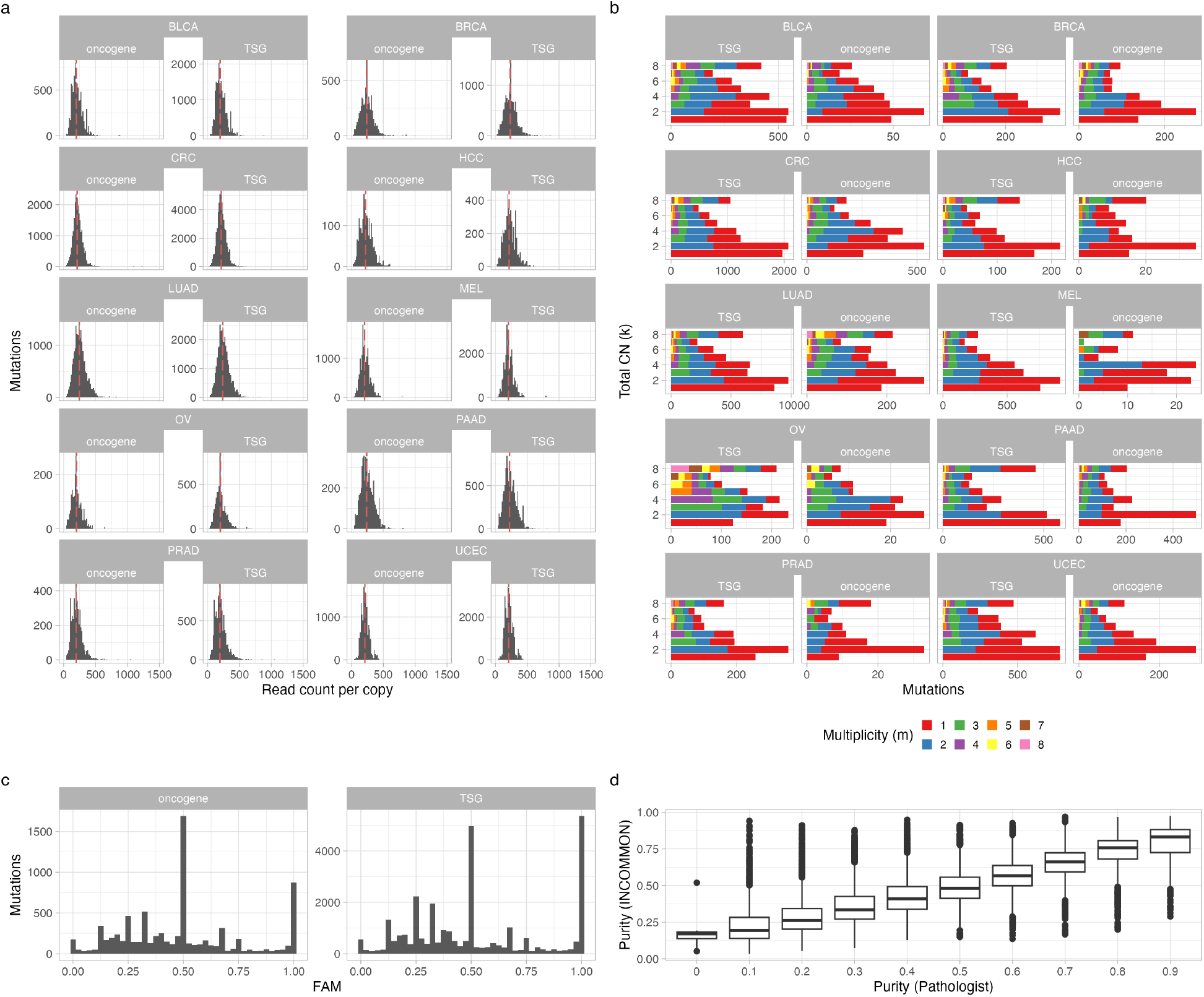
a. Distribution of maximum a posteriori (MAP) values of read count rate per copy *η*, across the main tumour types of the MSK-MET cohort. Red lines indicate median values. b. Distribution of MAP values of total copy number *k* and multiplicity *m*. c. Distribution of the MAP values of FAM. d. Comparison of MAP values of the sample purities with the pathologists’ estimates.

**Supplementary Figure S12.**
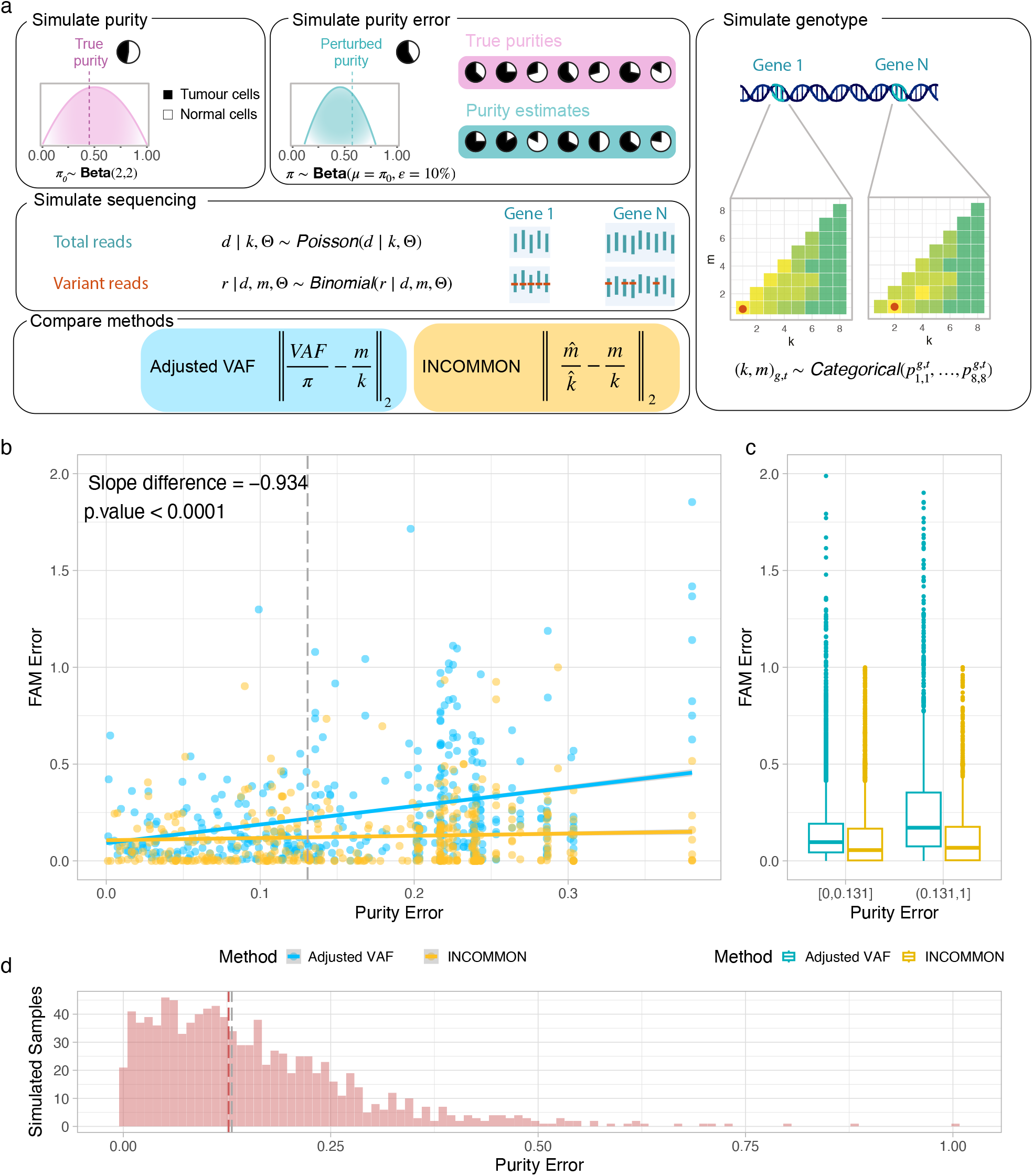
a. Simulation framework used to compare INCOMMON and adjusted VAF approaches: tumour purities were sampled from a Beta distribution, then perturbed to introduce random estimation error. Mutation copy number (k) and multiplicity (m) for randomly sampled mutant genes were sampled from empirical prior distributions, followed by simulation of sequencing read counts. Estimated FAM values from both methods were compared against the ground truth. b,c. Adjusted VAF exhibits substantially larger error than INCOMMON as the purity error increases. d. Distribution of simulated purity errors, with median value (red line) matching that of TCGA consensus purity estimates (grey line).

**Supplementary Figure S13.**
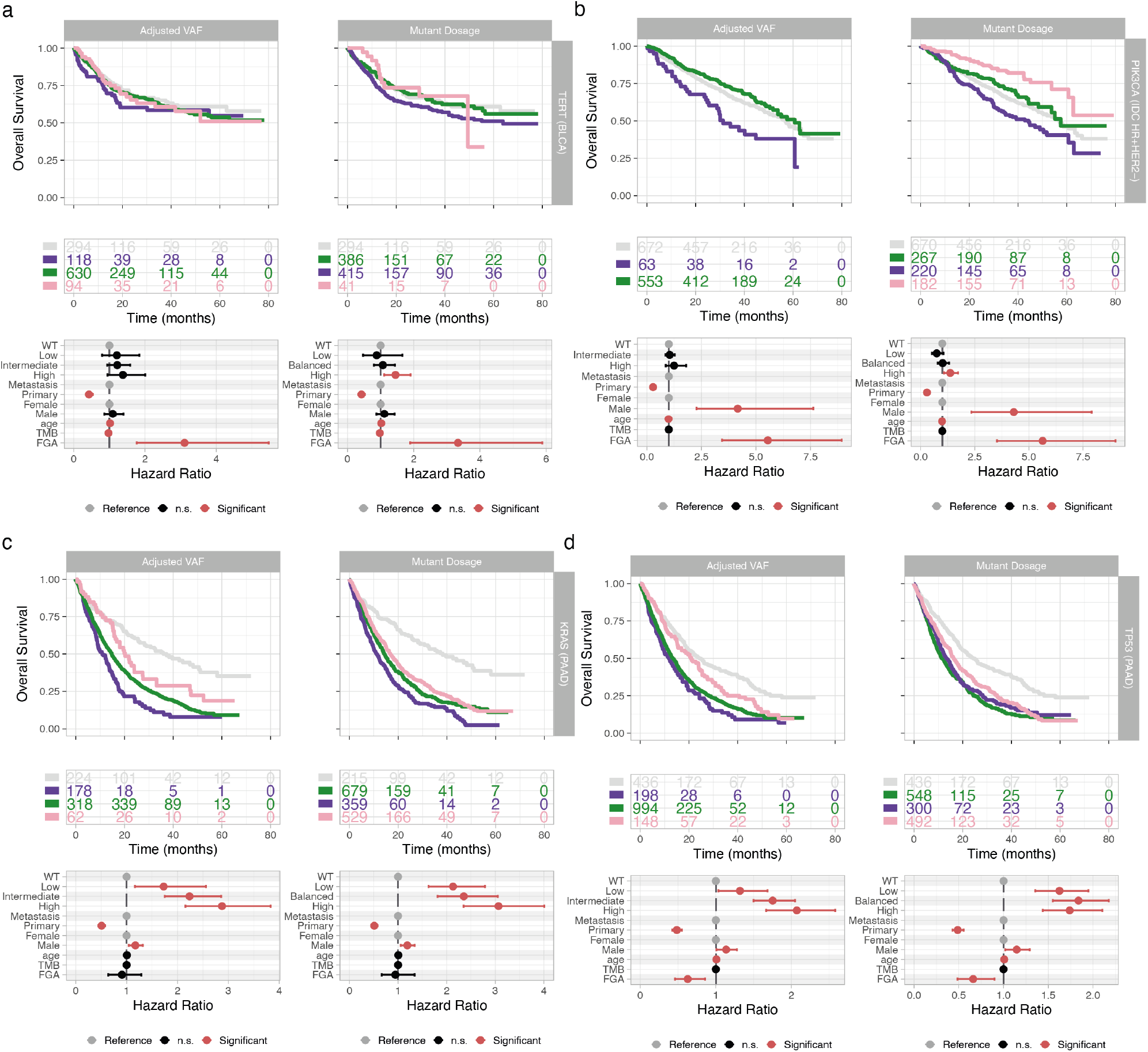
Representative examples comparing Mutant Dosage and Adjusted VAF stratifications for overall survival (OS). Both approaches define three classes (Low, Intermediate/Balanced, High) and are evaluated using Kaplan–Meier curves and multivariate Cox regression adjusted for primary vs. metastatic status, sex, age, TMB, and FGA. a–b. Differences in prognostic marker discovery rates arise because Adjusted VAF generally mirrors the trend of decreasing OS with increasing Mutant Dosage, but often fails to reach statistical significance due to higher uncertainty in hazard ratio (HR) estimates. Examples include TERT-mutant bladder cancer and PIK3CA-mutant HR+/HER2-breast cancer. c. In most settings, both methods identify similar prognostic groups with comparable HR estimates, although Adjusted VAF typically produces wider confidence intervals; KRAS-mutant pancreatic cancer illustrates this case. d. In a smaller set of cases, adjusted VAF yields the same number of classes but assigns more distinct and interpretable HR values across groups than Mutant Dosage; TP53-mutant pancreatic adenocarcinoma is an example.

**Supplementary Figure S14.**
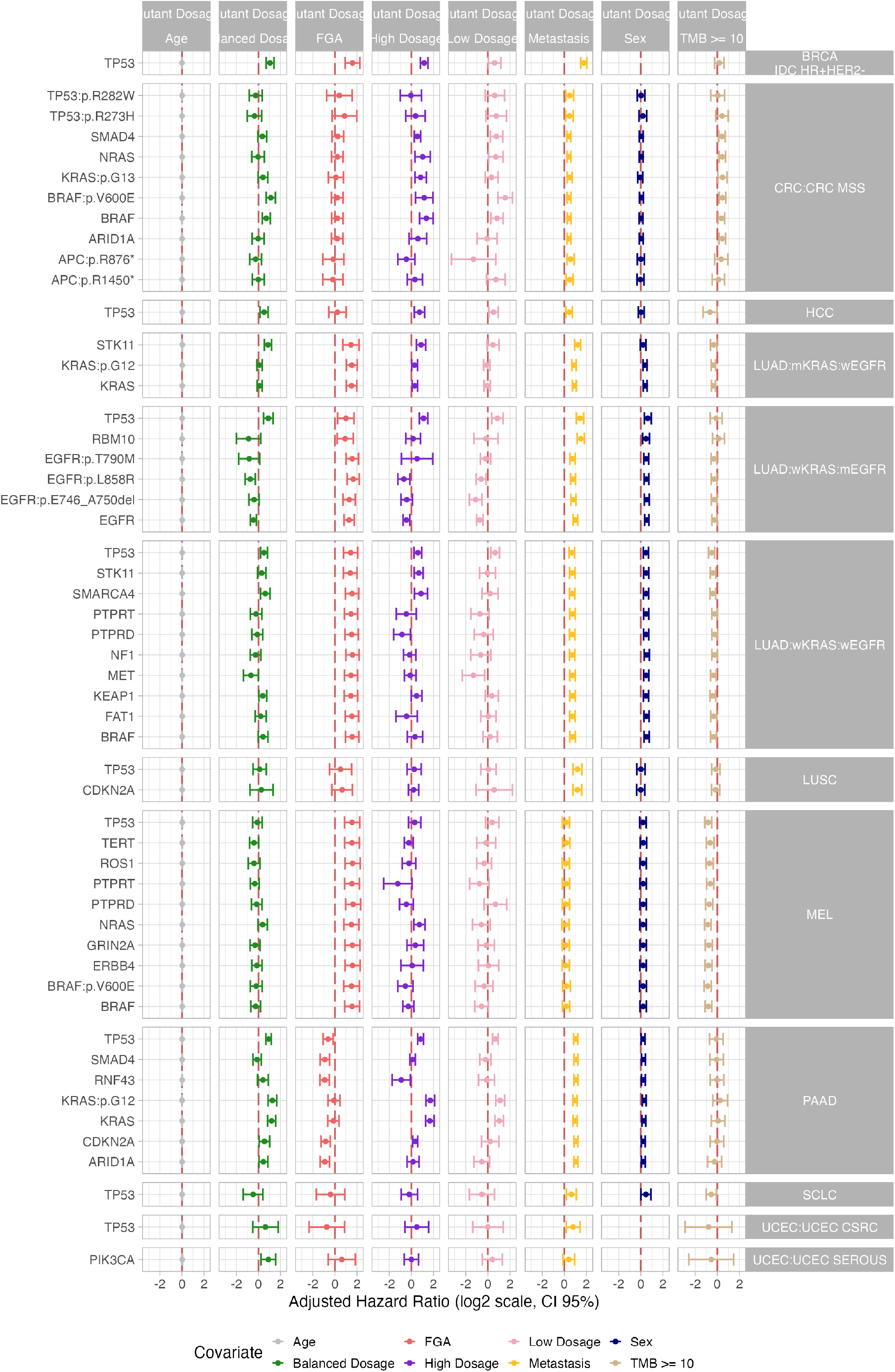
Adjusted hazard ratios (HR, log2 scale) with 95% confidence interval for all the covariates included in the multivariate Cox regression models of overall survival. The included covariates are mutant dosage class, age, sex, TMB, FGA and sample type. Red vertical lines indicate HR = 1. For each tumour type, only the most frequently mutated genes are shown.

**Supplementary Figure S15.**
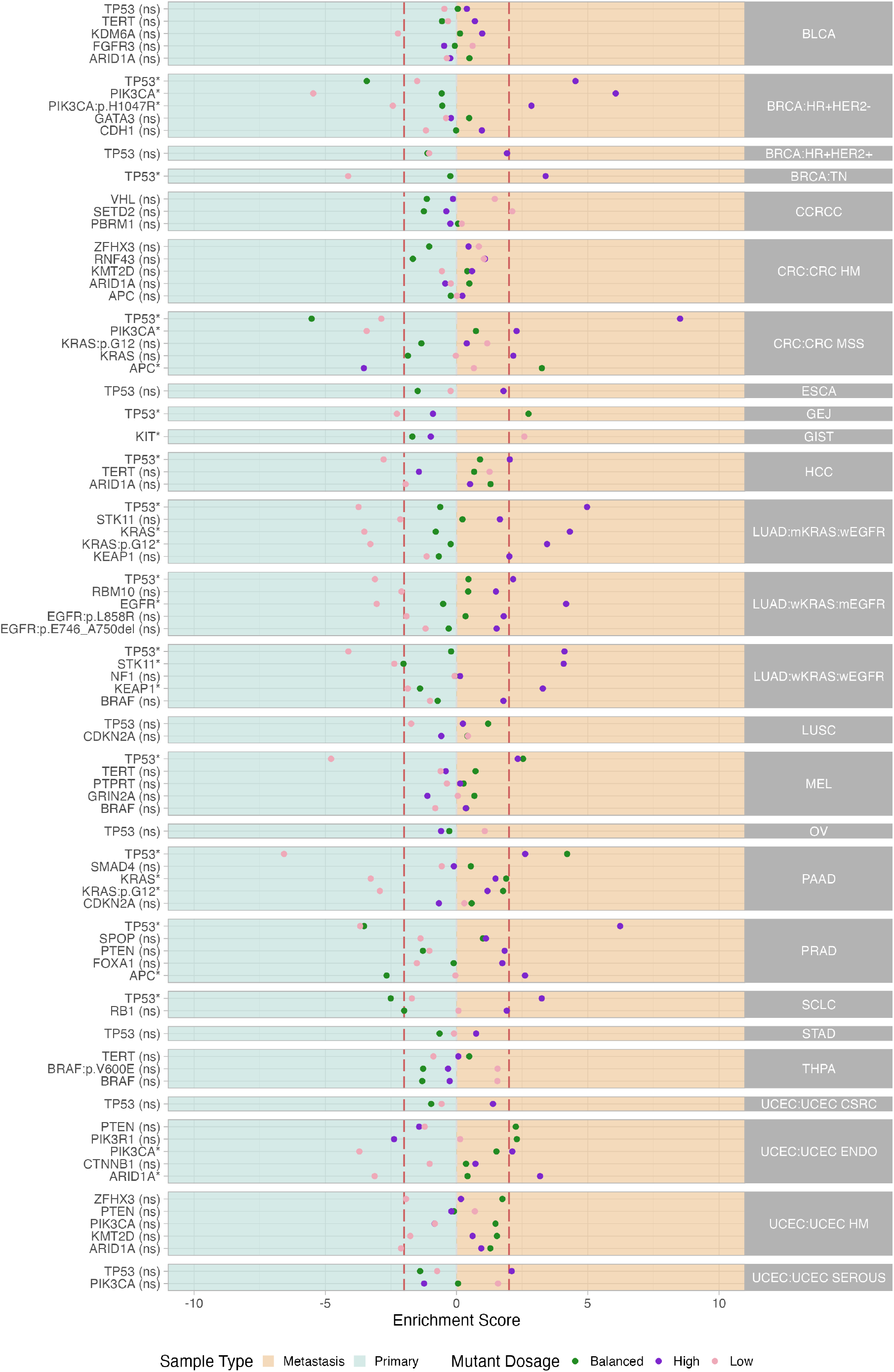
Enrichment scores *r* in primary tumours vs metastases of low, balanced and high mutant dosage classes, across the most frequently mutated genes per tumour type in the MSK-MET cohort. The enrichment score is evaluated through a two-tailed Fisher’s exact test with a significance level *α* = 0.1 and using standardised residuals. Vertical dashed lines indicate thresholds of large effect size (*r* = 2).

**Supplementary Figure S16.**
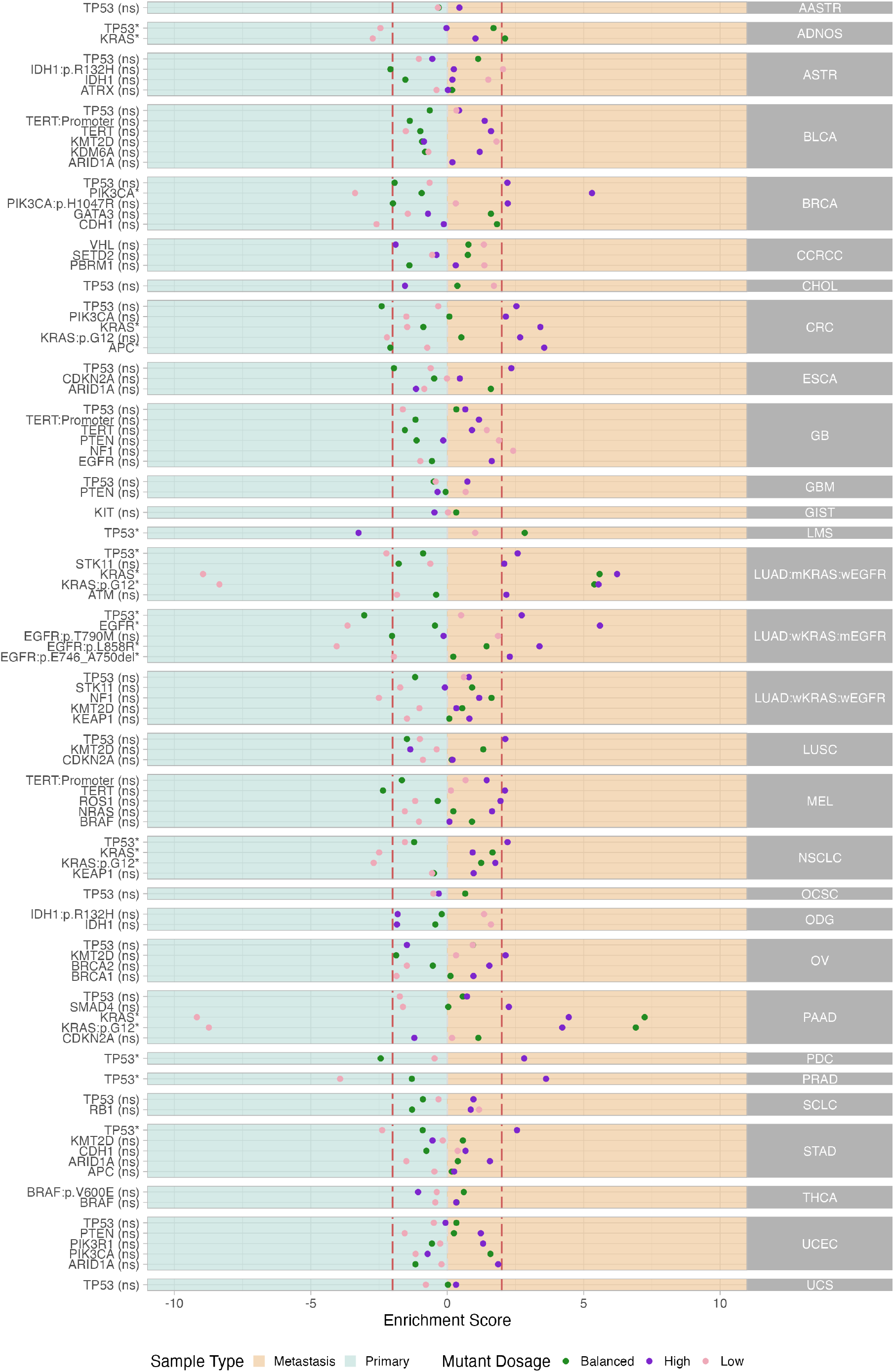
Enrichment scores *r* in primary tumours vs metastases of low, balanced and high mutant dosage classes, across the most frequently mutated genes per tumour type in the GENIE-DFCI cohort. The enrichment score is evaluated through a two-tailed Fisher’s exact test with a significance level *α* = 0.1 and using standardised residuals. Vertical dashed lines indicate thresholds of large effect size (*r* = 2).

**Supplementary Figure S17.**
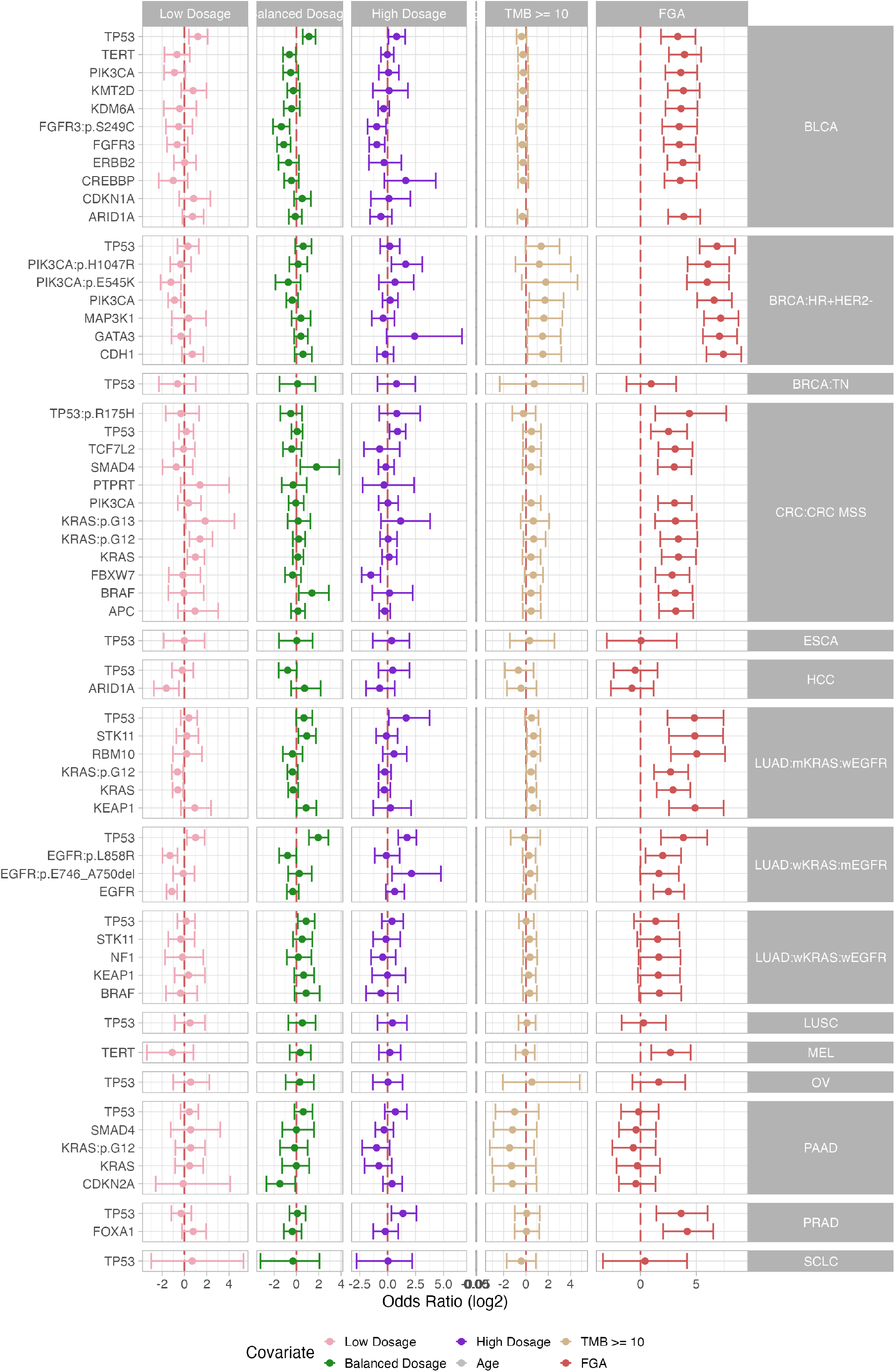
Adjusted OR of metastatisation (log2 scale) with 95% confidence interval, from multivariate logistic regression models. Covariates include low, balanced and high mutant dosage classes, age, tumour mutational burden (TMB) and fragment of genome altered (FGA). Red vertical lines indicate OR = 1. For each tumour type, only the most frequently mutated genes are shown.

**Supplementary Figure S18.**
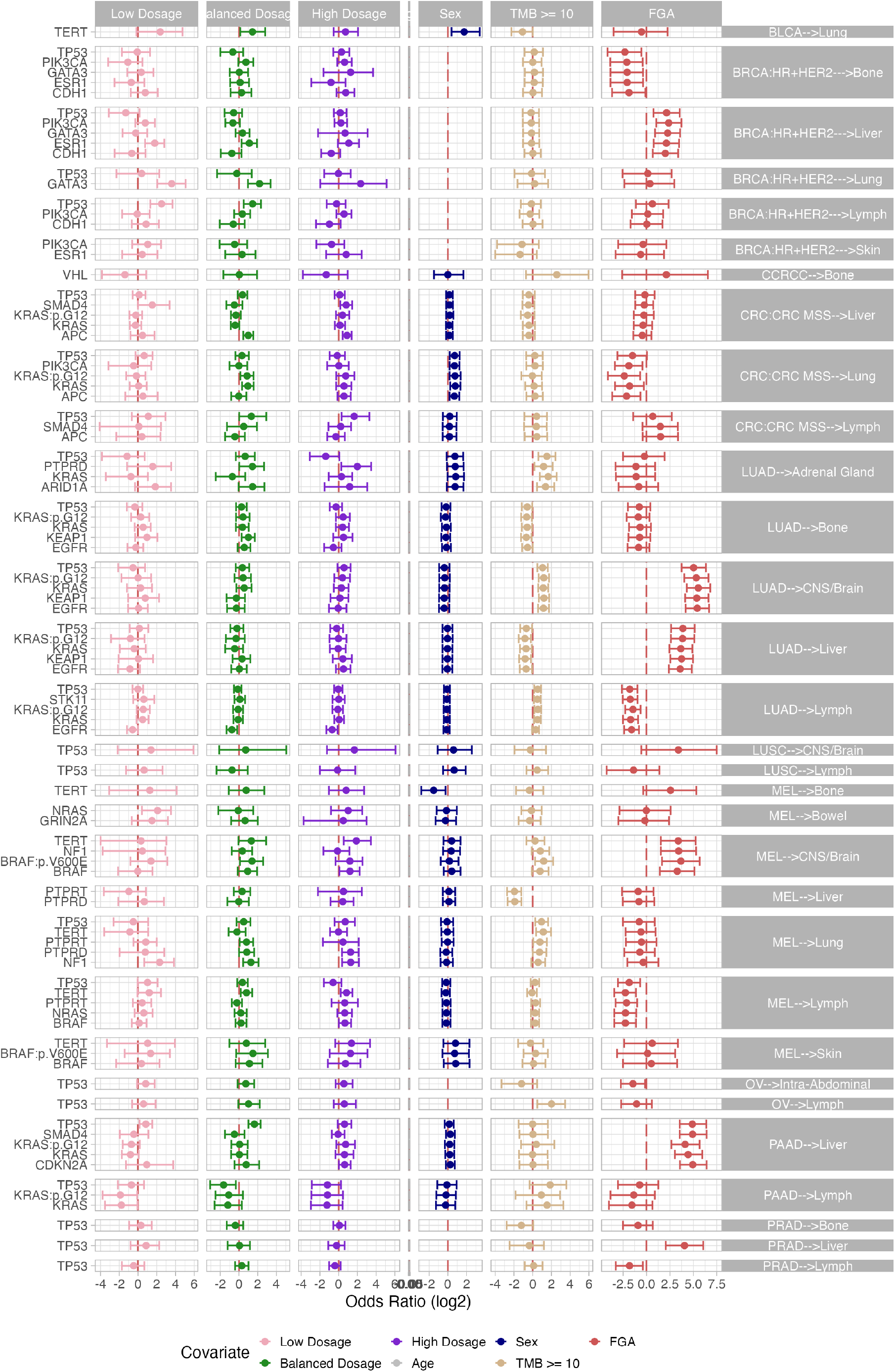
Adjusted OR of organotropic trajectories (log2 scale) with 95% confidence interval, from multivariate logistic regression models. Covariates include low, balanced and high mutant dosage classes, age, sex, tumour mutational burden (TMB) and fragment of genome altered (FGA). Red vertical lines indicate OR = 1. For each tumour type, only the most frequently mutated genes are shown.

**Supplementary Figure S19.**
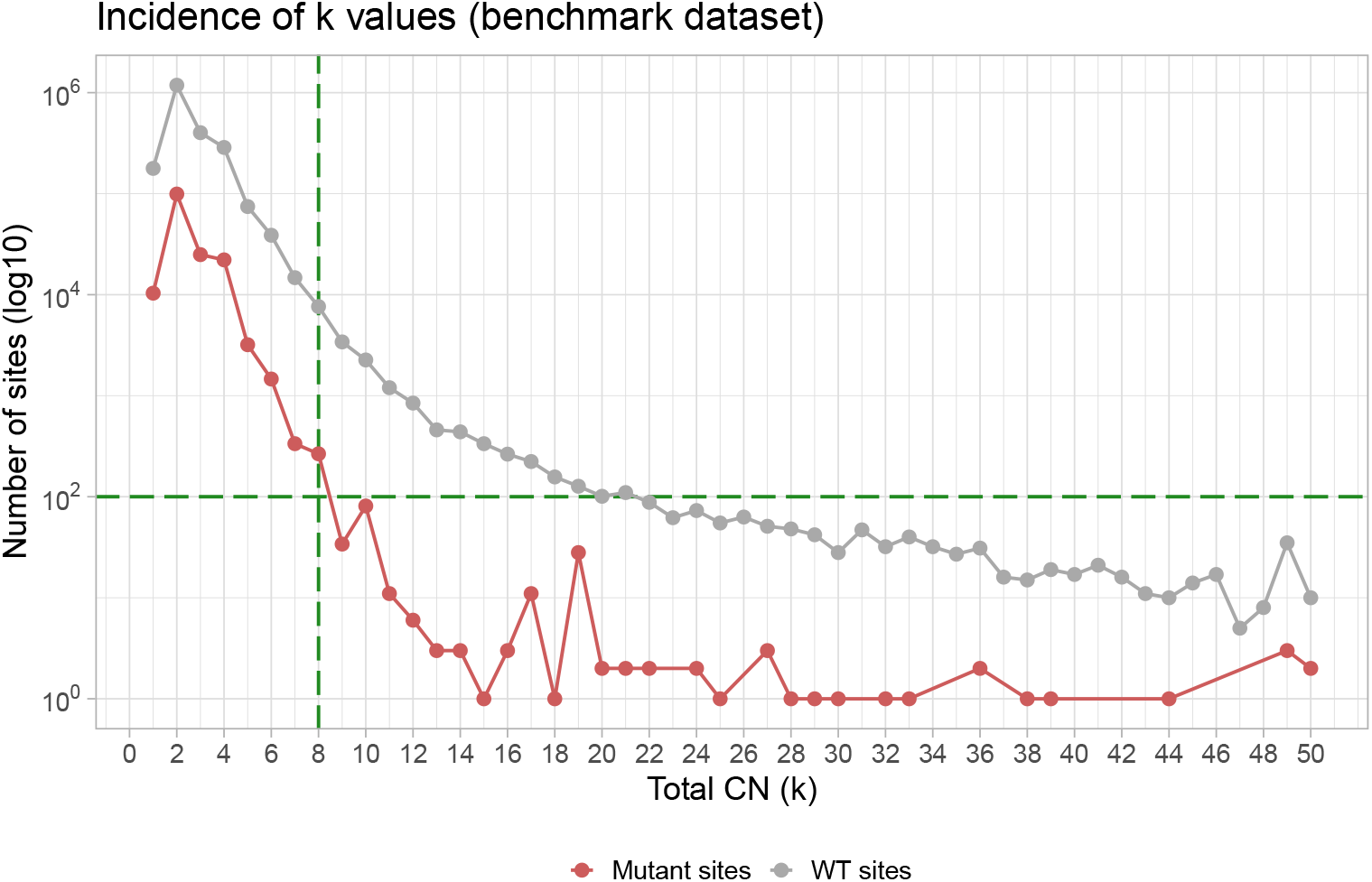
Number of mutant (red) and wild-type (gray) sites per total copy number across 8,339 samples from PCAWG, HMF and TCGA. In the analyses reported in this work, we set the maximum total copy number at *k*_*max*_ = 8 to reduce the computational burden. High copy gains were more frequent in wild-type regions, and for copy number values *k* > *k*_*max*_ we observed a significant drop in the number of mutant sites per copy-number (less than 100).

